# Somatic mutations in Alzheimer-associated tetraploid neurons

**DOI:** 10.1101/2024.01.13.24301214

**Authors:** Noelia López-Sánchez, Alberto Rábano, José M. Frade

**Affiliations:** Department of Molecular, Cellular and Developmental Neurobiology, Cajal Institute, IC-CSIC, Madrid, Spain; Tetraneuron, Valencia, Spain; Department of Neuropathology and Tissue Bank, Fundación CIEN, Instituto de Salud Carlos III, Madrid, Spain

**Keywords:** Oxidative stress response genes, DNA repair genes, Cancer-related genes, Two-hit hypothesis, Senescence-associated genes, Alzheimer-associated genes, Parkinson-associated genes, Molecular pathogenicity

## Abstract

An early pathological process affecting the brain of Alzheimer’s disease (AD) is the reactivation of the cell cycle in neurons followed by somatic neuronal tetraploidization (NT). NT also increases with age, and ageing has been shown to correlate with the accumulation of DNA somatic mutations. In this study, we have evaluated the presence of somatic mutations including single nucleotide variants (SNVs) and indels in genomic DNA from tetraploid neurons obtained from the parietal cortex of AD patients, compared with diploid neurons from control individuals and AD patients. Here we show that, in contrast to somatic indels, the proportion of somatic SNVs (sSNVs) significantly increases in the exome of tetraploid neurons, having increased levels of T to C (A to G) transitions, a type of mutation that is associated with oxidative stress. This finding correlates with the over-representation of sSNVs in genes involved in oxidative stress response and DNA repair, suggesting that these alterations exacerbate oxidative DNA damage in tetraploid neurons. sSNVs affecting cancer-related (CR) genes showed a greater molecular pathogenicity score compared with those from a random sample of genes. We propose that neuronal tetraploidy is stochastically triggered through a CR mechanism in neurons whose DNA repair genes become mutated in an oxidative stress scenario. This mechanism likely participates in the etiology of AD.

## Introduction

Alzheimer’s disease (AD) is the most prominent cause of dementia and a major health threat as it affects a substantial proportion of elderly in western countries. One of the earliest pathological events observed in AD is cell cycle reentry in neurons [15], which may be followed by DNA synthesis. This results in neuronal hyperploidy (including full DNA replication generating somatic tetraploid neurons) [1, 32, 40, 59] as well as specific death of hyperploid neurons at later stages [1, 4]. Compelling evidence indicates that neuronal tetraploidy (NT) precedes and recapitulates AD [32], while NT prevention in the cerebral cortex of 5xFAD mice, a murine model of AD [45], correlates with enhanced cognition [33, 34]. NT can affect neuronal structure and function [14], and triggers synaptic dysfunction [4], which has been postulated to disturb proper functioning of neural networks leading to cognitive deficits [3]. Furthermore, forced cell cycle reentry in neurons results in classical hallmarks of AD, including tau protein hyperphosphorylation and neurofibrillary tangles (NFT), increased APP processing and extracellular deposits of Aβ, delayed neuronal cell death, gliosis, and cognitive deficits [23, 35, 48, 49]. Musi and collaborators [41] have shown that genes involved in cell cycle progression are upregulated in human NFT-containing neurons. Moreover, a detailed proteomic analysis performed in six different brain regions from AD patients, as compared with control individuals, has identified several signaling pathways involved in cell cycle regulation as being widely dysregulated in severely affected regions of the AD brain [58]. Altogether, this evidence indicates that NT participates in the etiology of AD.

The mechanism leading to neuronal cell cycle reentry and somatic neuronal tetraploidization (SNT) in AD is currently unknown. Since aging is the main risk factor for sporadic AD [18], we hypothesized that the molecular changes associated with aging likely participate in the generation of AD-associated NT. Besides large genome structural variants, two types of small somatic mutations can accumulate during cell aging: single nucleotide variants (SNVs) and short insertions or deletions (indels) [54]. We therefore focused on the analysis of somatic DNA mutations that accumulate during aging in human neurons [31], resembling the progressive increase of somatic tetraploid neurons that precede the neuropathological hallmarks of AD [32]. To study the connection between somatic DNA mutations and NT, we performed whole exome sequencing from diploid and tetraploid neurons isolated from the parietal cortex of AD patients as well as diploid neurons from control individuals. This analysis demonstrated that an increase of oxidative stress-associated somatic mutations, mostly C>T transitions, can be found in tetraploid neurons from AD patients as compared to diploid (AD) or control (control) neurons. This effect is associated with enhanced accumulation of somatic mutations in genes regulating oxidative stress-response and DNA repair. The high pathogenicity of the somatic mutations observed in cancer-related (CR) genes suggests that somatic neuronal tetraploidization can be assimilated to an oncogenic-like process, as previously suggested by Zhu and collaborators [63, 64].

## Materials and Methods

### Human samples

Samples of parietal cortex from Alzheimer’s disease patients (n=3) and from healthy controls (n=3) were provided by the Banco de Tejidos Fundación CIEN (Madrid, Spain) and the Banco de Cerebros de la Región de Murcia (Hospital Virgen de la Arrixaca, Murcia, Spain). Immediately after post mortem brain extraction, each brain was sagittally cut, and slices from the right brain half were frozen in -60°C isopentane. A full neuropathological examination of each brain was performed on the left brain half, and samples from either Braak 0 (control) or Braak V-VI patients were chosen for this analysis (Additional file 1). Severity of Alzheimer pathology was scored according to the National Institute on Aging – Alzheimer’s Association Guidelines, following the “ABC” protocol [39]. Consequently, total amyloid burden (“A” score) was determined according to the Thal staging system; the stage of neurofibrillary pathology was established according to the Braak (“B” score) scheme; and the frequency of neuritic plaques in associative cortex according to the CERAD protocol (“C” score) was registered. All control cases were free from significant neurodegenerative or cerebrovascular pathology. Two of the AD cases (identification codes AD166 and AD409) displayed, additionally, moderate cerebrovascular pathology, while one of them (identification code AD423) showed high intensity small-vessel disease, Lewy-type pathology limited to the olfactory bulb and TDP-43(+) pathology (Limbic-predominant age-related TDP-43 encephalopathy, LATE) limited to the medial temporal lobe (stage 2). The identification codes of the AD patients and control individuals were not known to anyone outside the research group.

### Antibodies

The rabbit anti-NeuN polyclonal antibody (Merck Millipore) and the mouse anti-NeuN monoclonal antibody (clone A60, Merck Millipore) were both diluted 1/800 for flow cytometry. Secondary anti-mouse or anti-rabbit antibodies, generated respectively in donkey or goat and conjugated to Alexa Fluor 488, were purchased from Invitrogen. These antibodies were used at 1/400 dilution for cytometry.

### Cell nuclei isolation and immunostaining

Cell nuclei isolation was performed following a procedure described previously [32]. Briefly, a cube of 5-8 mm edge of fresh-frozen human parietal cortex was placed in 3.0 ml ice-cold, DNase-free PBS containing 0.1 % Triton X-100 (Sigma-Aldrich) (PTx) containing protease inhibitor cocktail (Roche). Cell nuclei were then isolated by mechanical disaggregation using a dounce homogenizer and then diluted up to 4.5 ml with PTx. Undissociated tissue was removed by centrifugation in 1.5-ml minifuge tubes at 200xg for 1.5 min at 4°C. Supernatants were reserved, and pellets were washed with 1.5 ml PTx and centrifuged at 100xg for 2 min at 4°C. Supernatants were collected and added to the previous ones. These samples were 8-fold diluted with PTx and centrifuged at 400xg for 4 min at 4°C. Supernatants, which lacked cell nuclei, were discarded, and the pellet incubated at 4°C in 800-1,000 μl cold PTx for at least 10 min, prior to mechanical disaggregation by gently swirl of the vial. The quality and purity of the isolated nuclei was checked microscopically after staining with 100 ng/ml DAPI.

Nuclear immunostaining with anti-NeuN antibodies was performed as previously described [32]. Briefly, primary and secondary antibodies were added to isolated unfixed nuclei containing 5 % of fetal calf serum (FCS) and 1.25 mg/ml of BSA. In control samples, the primary antibodies were excluded. Finally, the reaction was incubated O/N at 4°C in the dark. Immunostained nuclei were filtered through a 40-μm nylon filter, able to retain big aggregates but not nuclei. Then, the volume was adjusted to 800-1000 μl with DNase-free PTx containing 40 μg/ml PI (Sigma) and 25 μg/ml DNAse-free RNAse I (Sigma), and incubated for 30 min at RT. The quality of the nuclei and specificity of immunostaining signal was checked by fluorescence microscopy.

### FACS

Flow cytometry was carried out as described previously [32], using a FACSAria cytometer (BD Biosciences, San Diego, CA) equipped with a 488-nm Coherent Sapphire solid state and 633-nm JDS Uniphase HeNe air-cooled laser. The emission filters used were BP 530/30 for Alexa 488, BP 616/23 for PI. Data were analyzed with FACSDiva (BD Biosciences) and Weasel 3.0.1 (Walter and Eliza Hall Institute of Medical Research) softwares, and displayed using biexponential scaling. Cellular debris, which was clearly differentiated from nuclei due to its inability to incorporate PI, was gated and excluded from the analysis. DNA content histograms were generated excluding doublets and clumps by gating on the DNA pulse area versus its corresponding pulse height. For maximum doublet resolution, minimal flow rate was used in all experiments. The exclusion of doublets was confirmed by checking the DNA pulse area versus the pulse width of the selected population. Two rounds of cell sorting were performed to get a population of NeuN-positive nuclei enriched in tetraploid neurons (4C samples). Then, an aliquot of each 4C sample was used to estimate the proportion of tetraploid nuclei through flow cytometry, quantified by using the BD FACSDiva™ Software. The final proportion of tetraploid neurons in the 4C samples was above 70 % except for the individual AD166, which was discarded for the analysis.

### Whole genome amplification (WGA)

Genomic DNA from 430-550 diploid or tetraploid neuronal nuclei was isolated and amplified using the PicoPLEX WGA kit (New England Biolabs). WGA amplification included one cycle of 95°C (2 min), followed by 16 cycles of 95 °C (15 s), 15 °C (50 s), 25 °C (40 s), 35 °C (30 s), 65 °C (40 s), and 75 °C (40 s). Successful amplification was confirmed by agarose gel electrophoresis. DNA from PicoPLEX was purified using the Gene elute PCR clean-up kit (Sigma Aldrich). Cleaned-up products were quantified using a Nanodrop spectrophotometer (Thermo Scientific).

### Libraries preparation, exome sequencing, and bioinformatics analysis

Libraries were prepared using Comprehensive Exome Panel technology (Twist Bioscience, South San Francisco, CA, United States) and sequenced on NovaSeq 6000 System™ (Illumina, San Diego, CA, United States). The raw FASTQ files generated were first evaluated using FastQC (https://www.bioinformatics.babraham.ac.uk/projects/fastqc) and Trimmomatic [6] to remove bases, adapters, and other low-quality sequences. Each FASTQ file was then aligned against the GRCh38/hg38 version of the reference human genome by BWA-mem [25]. BAM files were generated with SAMtools [26], and the elimination of optical and PCR duplicates was carried out with Sambamba [53]. SNVs and indels were identified using VarScan 2 [22]. Variants were annotated with information from the following databases containing functional information (Ensembl, Pfam), population information (dbSNP, 1000 Genomes, ESP6500, ExAC, gnomAD), in silico prediction of functional impact (dbNSFP, dbscSNV), cancer related information (COSMIC, ICGC), and clinical and phenotypic trait information (ClinVar, OMIM, HPO). The tetraploid neuron-specific variants in each AD patient were identified as those with null frequency in the diploid sample and frequency > 0 in the tetraploid one. All the statistical analyses of the results were made with R software v4.1.2. In those cases where conditions were compared the Student’s t test was used with p-value < 0.05 as significant threshold. SNVs and INDELS were analyzed separately. Both, the percentage of variants per chromosome and the number of variants per chromosome size, were calculated among samples and conditions. The density and frequency of these changes were represented across each chromosome. In the case of SNVs variants, the number and frequency of each type of change were also analyzed. Likewise, the same analysis was performed pre-selecting those variants affecting genes related with Alzheimer’s disease- and Parkinson’s disease (based on literature search), Cancer (based on 409 proto-oncogenes and tumor suppressor genes that are used for diagnosis of human cancer [OncoNIM Seq409 diagnostic system (NIM Genetics)]), and DNA repair, response to oxidative stress, senescence, and cancer (based of gene ontology search) (see Additional file 2 for a list of the respective genes that were analyzed per group). Molecular pathogenicity of proteins was evaluated using the DG prediction score (Dreamgenics; https://www.dreamgenics.com), calculated from the dbNSFP database [28–30] that brings together the *in silico* predictions of twelve different algorithms: ALoFT [2], DEOGEN2 [50], FATHMM [52], FATHMM-MKL (https://fathmm.biocompute.org.uk), FATHMM-XF (https://fathmm.biocompute.org.uk/), META-LR [13], META-SVM [13], SIFT [20], PROVEAN [9], Mutation Assesor [17], Mutation Taster [62], and LRT [10].

## Results

### Increase of SNVs, but not indels, in tetraploid neurons from AD patients

The absolute number of mutations, including SNVs and indels, was quantified in genomic DNA from tetraploid neurons obtained from the parietal cortex of AD patients, compared with diploid neurons from control individuals (NoAD) and AD patients. To this aim, whole genomic DNAs from 430-550 neuronal (i.e. NeuN^+^) nuclei per individual were whole-genome amplified (Additional file 1) and subjected to deep next-generation sequencing, as described in the methodological section. This analysis demonstrated that AD-derived tetraploid neurons contain a statistically-significant increase (16 %) of mutations when compared with diploid neurons from AD patients (Fig. 1a). In contrast, no significant differences were observed between diploid neurons from AD patients compared to NoAD individuals, indicating absence of age bias in our analysis, as previously reported by Li and collaborators [27]. These mutations in AD-derived tetraploid neurons affected similar chromosomal regions as those detected in diploid neurons from NoAD and AD patients (see circos plots in Fig. 1b), suggesting the existence of specific mutation hot spots in the genome of human neurons as in other cell types, likely due to aberrant repair of breaks in sequence-specific contexts [43]. The observed increase of mutations in tetraploid neurons was specific for SNVs, representing a 17 % increase when compared with diploid neurons from AD patients (Fig. 1c), since the absolute number of indels was similar in all neuronal types (Fig. 1d).

**Fig. 1.**
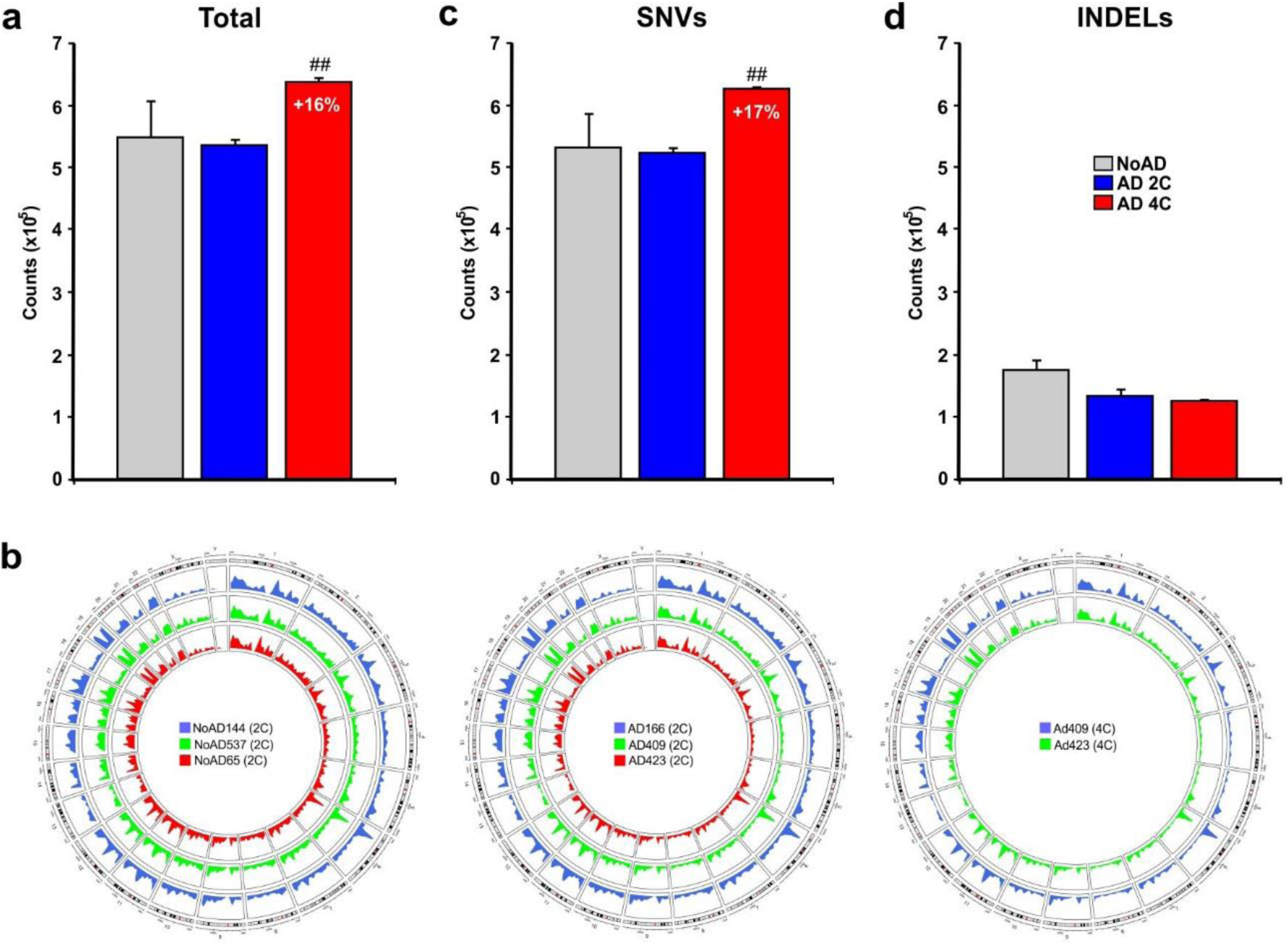
Total mutation counts in diploid neurons from control (noAD) and AD patients (AD 2C) as well as in tetraploid neurons from AD patients (AD 4C). **a** Total somatic mutations observed in noAD diploid neurons (grey), AD diploid neurons (blue) and AD tetraploid neurons (red). **b** Total sSNVs observed in noAD diploid neurons (grey), AD diploid neurons (blue) and AD tetraploid neurons (red). **c** Total indels observed in noAD diploid neurons (grey), AD diploid neurons (blue) and AD tetraploid neurons (red). **d** Circos plots illustrating the chromosomal distribution of somatic mutations observed in noAD diploid neurons, AD diploid neurons and AD tetraploid neurons from the individuals analyzed in this study. *p<0.05, **p<0.01 (Student’s *t* test).

### Specific increase of de novo-generated SNVs in tetraploid neurons from AD patients

We determined the distribution of frequencies (f) at which the different SNVs were present in the genomic DNAs. This analysis revealed one major peak of SNVs with low frequency (f < 0.1) and two minor peaks at f = 0.5 and f = 1.0 in the three neuronal populations (Fig. 2a). We interpreted these latter two peaks as germline SNVs (gSNVs) (or allelic variants) in heterozygosity and homozygosity, respectively. In contrast, the major peak with f < 0.1 would represent somatic SNVs (sSNVs) generated *de novo* as the brain ages [31] and/or when it undergoes neurodegeneration [36, 46, 47]. This interpretation is consistent with the results obtained from the analysis of SNVs within the same AD individuals, classified as either common to both diploid and tetraploid neurons (i.e. germline variants) or specific for either diploid or tetraploid neurons (i.e. somatic mutations) (Additional file 3). Common SNVs showed conspicuous peaks with f = 0.5 and f = 1.0, plus an additional peak of SNVs at low frequency, likely derived from clonal somatic mutations during early development [37]. In contrast, SNVs unique for diploid and tetraploid were dramatically enriched for f < 0.1.

**Fig. 2.**
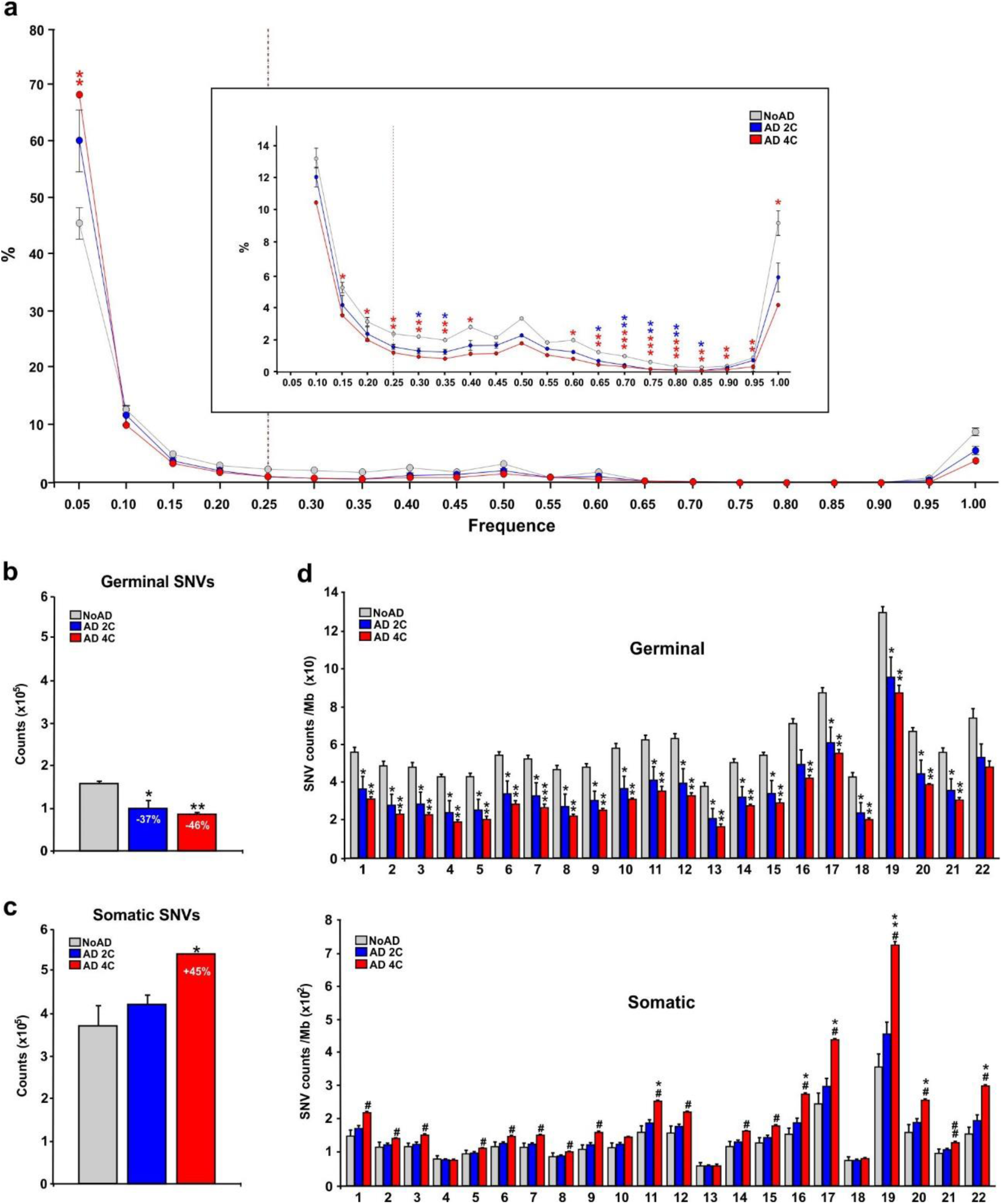
SNVs from germinal and somatic origin. **a** Relative frequency distribution of the SNVs in noAD diploid neurons (grey), AD diploid neurons (blue) and AD tetraploid neurons (red). Dotted line represent the threshold that was used to differentiate mutations from germinal (right) and somatic (left) origin. Box: zoom of the graph indicating percentages ranging from 0 to 14. **b** Germinal SNVs observed in noAD diploid neurons (grey), AD diploid neurons (blue) and AD tetraploid neurons (red). **c** Somatic SNVs observed in noAD diploid neurons (grey), AD diploid neurons (blue) and AD tetraploid neurons (red). **d** Germinal (left) and somatic (right) SNV counts per chromosomal Mb. Numbers identify the chromosomes. *p<0.05, **p<0.01; *p<005 (comparison NoAD vs AD 4C), ^#^p<0.05, ^##^p<0.01 (comparison NoAD vs AD 4C) (Student’s *t* test).

Based on these findings, we decided to set f = 0.25 as a threshold between the SNVs that are present at low frequency (f < 0.25), which would represent sSNVs, and those that appear at high frequency (f > 0.25), which would mostly be gSNVs. This setting is consistent with the presence of a substantial proportion of high frequency SNVs (i.e. gSNVs) in databases containing somatic nucleotide polymorphism, including the catalogue of somatic mutations in cancer (COSMIC; https://cancer.sanger.ac.uk/cosmic), the ICGC data portal (https://dcc.icgc.org/search/m), and the reference single nucleotide polymorphism database (dbSNP; https://www.ncbi.nlm.nih.gov/snp)], while most low frequency SNVs (i.e. sSNVs) were absent from these databases (Additional file 4).

As expected, the absolute number of gSNVs was similar in both diploid and tetraploid neurons from AD patients (Fig. 2b). Nevertheless, this number was significantly greater in noAD neurons (Fig. 2b), suggesting that a reduced proportion of gSNVs is linked to the genetic predisposition for the etiology of AD [5]. Interestingly, this analysis also indicated that the proportion of sSNVs becomes significantly increased (45 %) in tetraploid neurons from AD patients, as compared with diploid neurons from noAD individuals (Fig. 2c).

SNVs displayed differences among specific chromosomes (Fig 2d). sSNVs, especially those observed in tetraploid neurons, were enriched in chromosome 19, while three chromosomes (4, 13 and 18) showed low levels of sSNVs per Mb, which were equivalent in all neuronal types. This contrasts with the distribution of gSNVs, as these mutations showed decreased numbers per Mb in the AD-derived neurons independently of the chromosome that was analyzed.

To verify the specificity of the increase of sSNVs in tetraploid neurons, we performed a similar study on indels. Indels also showed one major peak with low frequency (f < 0.1) (i.e. somatic indels) and two minor peaks at f = 0.5 and f = 1.0 (germinal indels in heterozygosity and homozygosity, respectively) in the three neuronal populations (Additional file 5a). In addition, another peak of uncertain nature was also observed with f = 0.4. In all cases, the absolute number of germinal indels did not differ between diploid and tetraploid neurons from AD patients (Additional file 5b) but, as in the case of gSNVs, this number was increased in noAD neurons (Additional file 5b). In contrast, the number of somatic indels was similar in the three types of neurons analyzed (Additional file 5c), thus demonstrating that the increase of sSNVs observed in the tetraploid neurons (Fig. 2c) was specific for this latter type of mutation.

Quantitative differences between somatic indels and sSNVs were also observed when their chromosomal distribution was taken into consideration, further demonstrating the specificity of the observations for sSNVs. In contrast to sSNVs, which were significantly increased in most chromoses (Fig. 2d), the absolute number of somatic indels per Mb was either decreased or maintained in most chromosomes except for chromosome 19, in which a statistical significant increased number of somatic indels per Mb was observed (right panel in Additional file 5d). The analysis performed with germinal indels indicated that their chromosomal distribution was similar to that observed for gSNVs (left panel in Additional file 5d).

Overall, these findings support the hypothesis that NT is specifically associated with the increase of sSNVs (i.e. somatic mutations characterized by single nucleotide changes). These somatic mutations may participate in the generation of tetraploid neurons in the AD brain.

### Selective genome localization and specific function of sSNVs in tetraploid neurons from AD patients

The genomic region as well as the functional protein effect of the identified SNVs was studied, considering whether they are from germline or somatic origin. We found that sSNVs were enriched, as compared to gSNVs, in coding regions from all types of neurons (Additional file 6), being significantly reduced the proportion of sSNVs that are present in introns from tetraploid neurons (Additional file 7a). Moreover, all neurons from AD patients showed a significant increase of sSNVs located in 5’-untranslated regions (UTRs) (Additional file 7b), while the presence of these mutations in promoters was significantly increased in tetraploid neurons (Additional file 7b). gSNVs were slightly increased in coding regions (Additional file 6), exons (Additional file 7a) and promoters (Additional file 7b) from AD neurons, as compared to noAD neurons, suggesting that these mutations could participate in the known genetic predisposition of AD patients [5]. In accordance with the reduced proportion of sSNVs that are present in introns, tetraploid neurons contained significantly less sSNVs in intron acceptor sequences (Additional file 7c). These observations are consistent with a reduced functional effect on splicing of both germline and sSNVs, in AD neurons (Additional file 8a). A similar situation was found for stop-gaining mutations, which are dramatically decreased among sSNVs, in neurons from AD individuals, an effect that seems to be potentiated in tetraploid neurons (Additional file 8b). In addition, all types of neurons showed an enrichment of missense sSNVs, as compared to the proportion of missense gSNVs in these cells, an enrichment that is significantly enhanced in tetraploid neurons (Additional file 8c). Overall, these findings suggest that SNT is associated with sSNVs involved in protein malfunction due to missense mutations while largely maintaining intact the protein functional domains as both altered splicing and stop-gaining mutations become reduced.

### sSNVs from tetraploid neurons are enriched in T to C (T>C) transitions

Lodato and collaborators [31] demonstrated that somatic mutations, mostly T>C (or A>G in the DNA complementary strand) variants, accumulate in neurons during aging. We therefore identified the types of mutations that can be found in the studied neuronal types. This analysis indicated that T>C transitions are strongly enriched in tetraploid neurons, mostly within the pool of sSNVs (Fig. 3). This result was confirmed when SNVs common to diploid and tetraploid neurons and specific for either diploid or tetraploid neurons were studied. A dramatic increase of T>C transitions was specifically observed in SNVs specific for tetraploid neurons (Additional file 9).

**Fig. 3.**
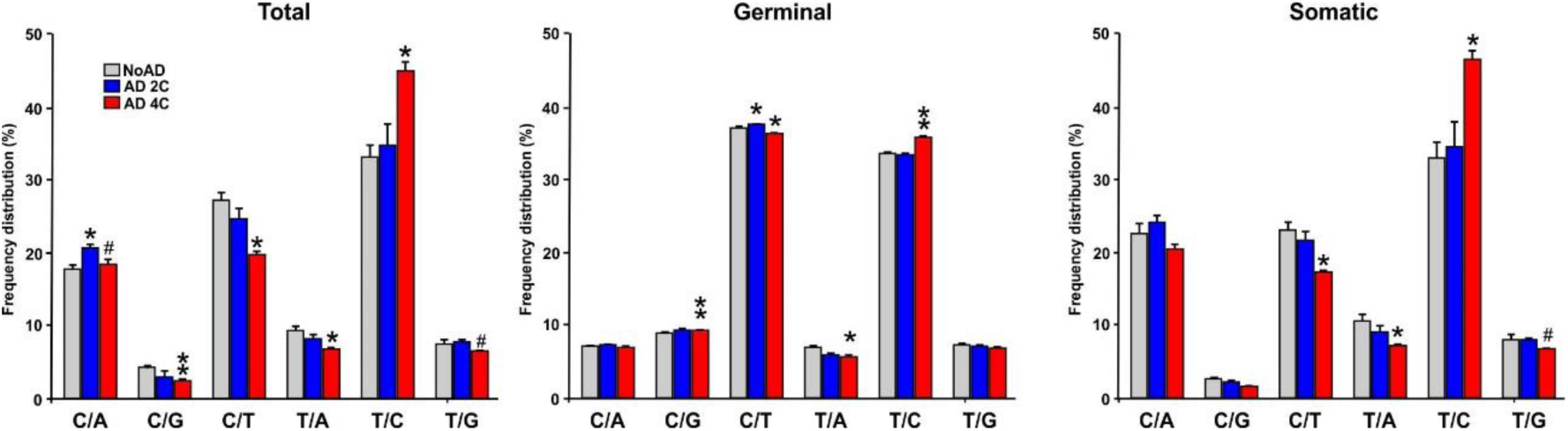
Percentage of the identified sSNVs in diploid neurons from noAD individuals and AD patients as well as in tetraploid neurons from AD patients. Percentage of the indicated sSNVs observed in noAD diploid neurons (grey), AD diploid neurons (blue) and AD tetraploid neurons (red). C/A (C>A, G>T), C/G (C>G, G>C), C/T (C>T, G>A), T/A (T>A, A>T), T/C (T>C, A>G), T/G (T>G, A>C). *p<0.05, **p<0.01 (comparison NoAD vs AD 4C), ^#^p<0.05 (comparison NoAD vs AD 4C) (Student’s *t* test).

### Enrichment of sSNVs in neurodegenerative disease-associated genes

Previous reports have pointed out that somatic variants in autosomal dominant genes known to be involved in AD can be associated with the etiology of this disease [44, 47]. We therefore explored whether sSNVs in AD risk genes (see list in Additional file 2) could be enriched in the tetraploid neurons. As a control, we used a list of Parkinson’s disease (PD)-related genes (see list in Additional file 2).

This analysis demonstrated that tetraploid neurons display a strong increase of total SNVs in AD-risk genes (27 % increase as compared to 16 % in the whole genome; Fig. 1a), showing a greater increase (59 % as compared to 45 % in the whole genome; Fig. 2c) when sSNVs are considered (Fig. 4a). This contrasts with the 40 % reduction of gSNVs that were present in the AD-risk genes from the tetraploid neurons (Fig. 4a).

**Fig. 4.**
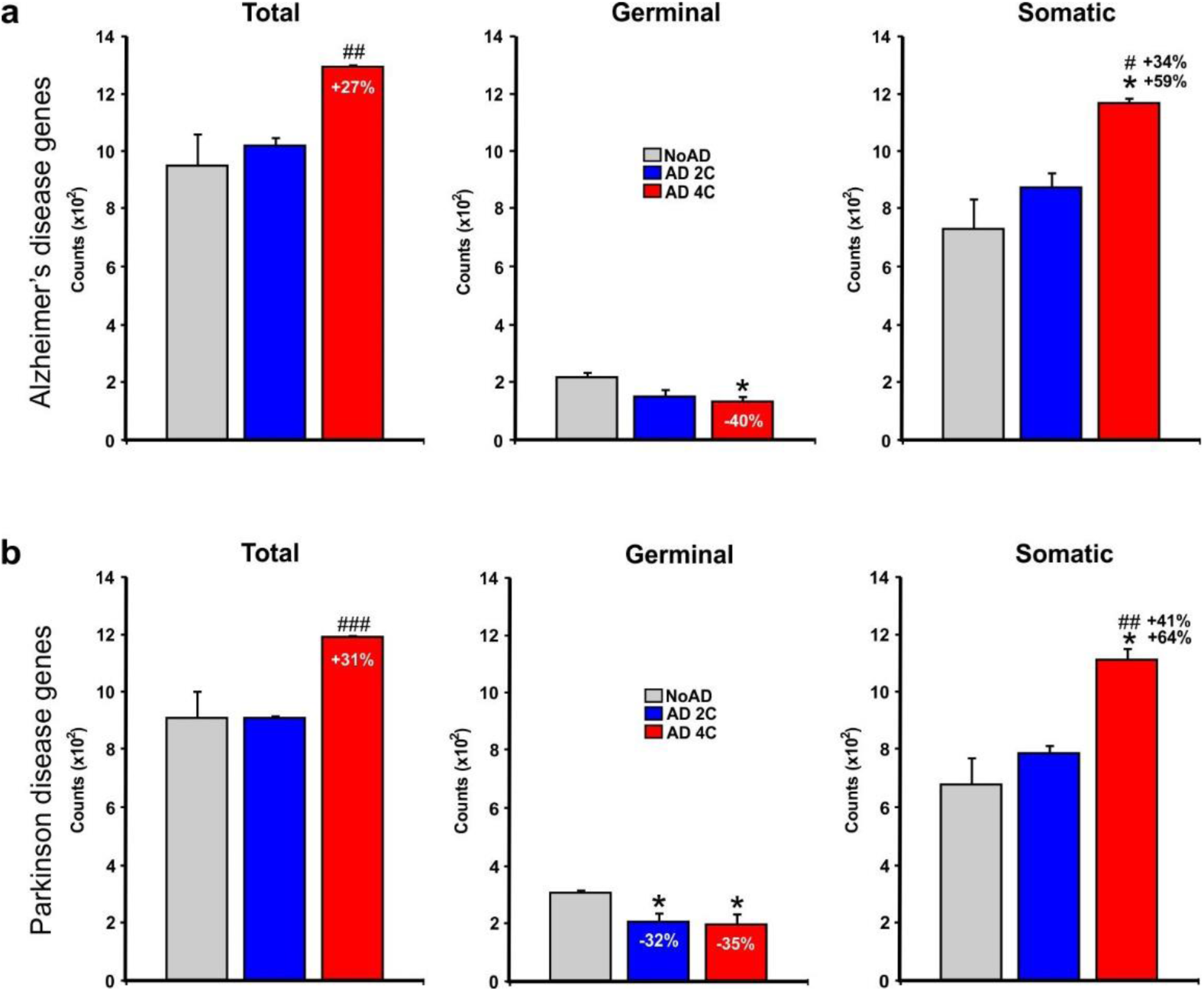
sSNVs counts in AD- and PD-risk gene sets of neurons isolated from noAD individuals and AD patients as well as in tetraploid neurons from AD patients. **a** Total (left), germinal (middle) and somatic (right) sSNVs observed in AD-risk genes from noAD diploid neurons (grey), AD diploid neurons (blue) and AD tetraploid neurons (red). **b** As is A for PD genes. *p<0.05, **p<0.01 (comparison NoAD vs AD 4C), ^#^p<0.05 (comparison NoAD vs AD 4C) (Student’s *t* test).

Interestingly, some differences could be detected when the AD-risk genes were considered individually. While ABCA7, APOE, BIN1, CD33, EPHA1, MAPT, PSEN2, SMARCA4, and PLD3 showed a significant increase in the number of sSNVs from tetraploid neurons, CD2AP and PSEN1 showed the opposite effect (Additional file 10).

Surprisingly, PD risk genes (Fig. 4b) showed also an increase of SNVs (31 %), which was enhanced in sSNVs (64 % increase), while gSNVs were also reduced (35 % in this case) as occurred with AD-risk genes.

### Enrichment of sSNVs in oxidative stress-associated genes

To begin to understand possible molecular mechanisms involved in AD-associated NT we focused on oxidative stress-associated genes (see Additional file 2 for a list of these genes), since oxidative stress is a known stressor participating in AD [8].

We found that tetraploid neurons accumulated SNVs in genes involved in antioxidant activity (38 %) (Fig. 5a), response to reactive oxygen species (ROS) (31 %) (Fig. 5b), hydrogen peroxide activity (29 %) (Fig. 5c), and oxidative stress (26 %) (Fig. 5d), while no statistically significant increase was detected in the proportion of SNVs affecting genes involved in both cell death by oxidative stress (Fig. 5e) and superoxide dismutase (SOD) activity (Fig. 5f) when tetraploid and diploid neurons were compared. This analysis also demonstrated a significant increase of sSNVs affecting a subgroup of oxidative stress-associated genes, higher than that observed in the global genome (45 %, see Fig. 2c). In this regard, a 64 % (Fig. 5a) and 61 % (Fig. 5b) of sSNVs were detected in genes involved in antioxidant activity and response to reactive oxygen species (ROS), respectively. In contrast, only 36 % (Fig. 5c) and 33 % increase (Fig. 5d) of sSNVs was detected in genes involved in hydrogen peroxide activity and oxidative stress, respectively; while no statistically significant increase was detected in the proportion of total sSNVs affecting genes involved in both cell death by oxidative stress (Fig. 5e) and superoxide dismutase (SOD) activity (Fig. 5f). Interestingly, with the exception of SOD genes, the presence of gSNVs in genes involved in oxidative stress was significantly reduced in the tetraploid neuron population (Fig. 5a-e).

**Fig. 5.**
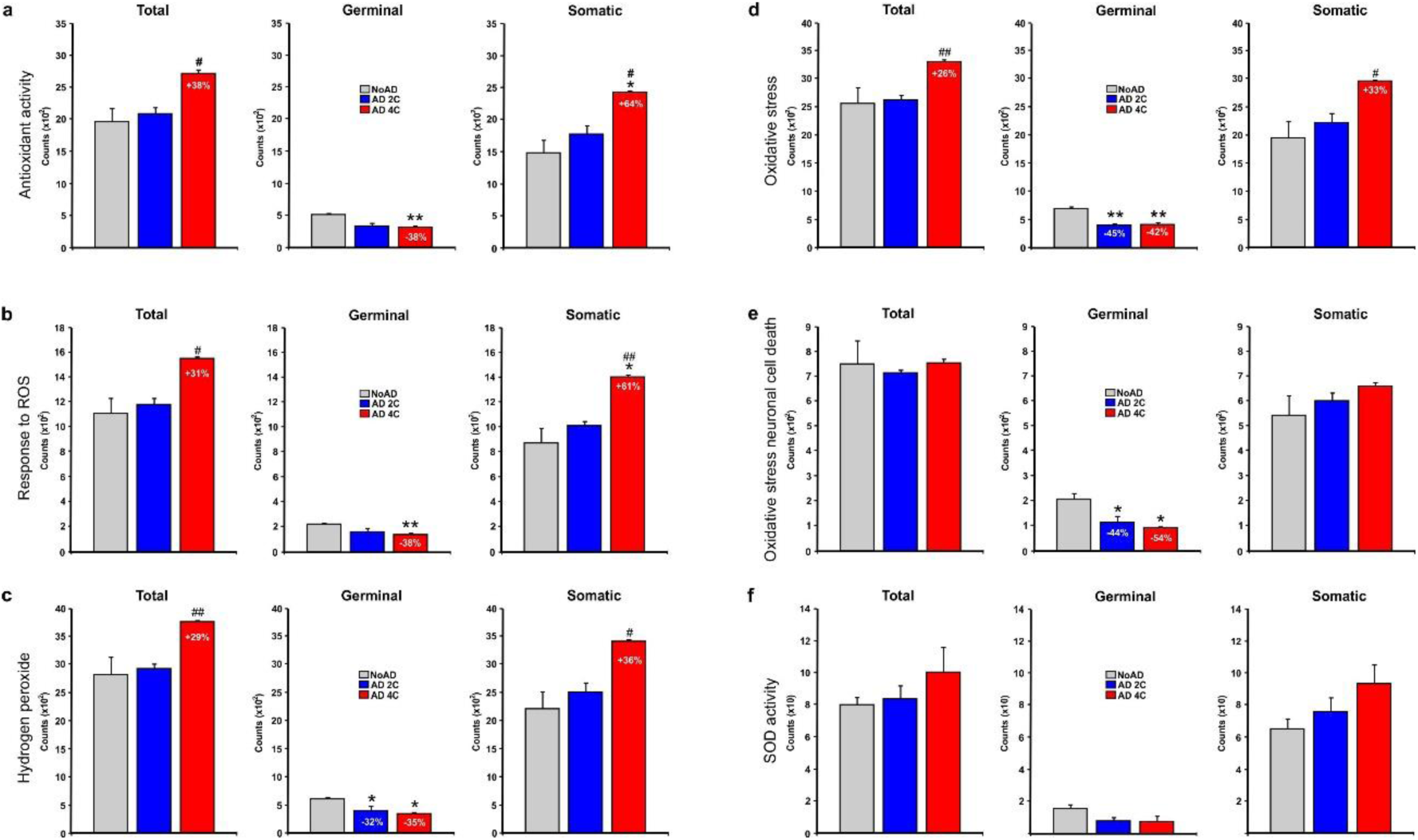
sSNVs counts in oxidative stress-associated gene sets of neurons isolated from noAD individuals and AD patients as well as in tetraploid neurons from AD patients. **a** Total (left), germinal (middle) and somatic (right) sSNVs observed in antioxidant activity genes from noAD diploid neurons (grey), AD diploid neurons (blue) and AD tetraploid neurons (red). **b** As is A for response to reactive oxygen species genes. **c** As is A for hydrogen peroxide genes. **d** As is A for oxidative stress genes. **e** As is A for oxidative stress neuronal cell death genes. f. As is A for SOD activity genes. *p<0.05, **p<0.01 (comparison NoAD vs AD 4C), ^#^p<0.05 (comparison NoAD vs AD 4C) (Student’s *t* test).

Overall, these findings indicate that oxidative stress-associated sSNVs involved in the response to oxidation and ROS accumulation likely participate in the generation of NT in the AD brain.

### Enrichment of sSNVs in DNA repair-associated genes

We found that SNVs become accumulated (31 % increase) in DNA repair genes (see list in Additional file 2) from tetraploid neurons (Fig. 6a), which represented a 64 % increase of sSNVs in comparison to a 45 % increase of sSNVs in the global genome (Fig. 2c). In contrast, gSNVs became decreased in these neurons by 35 % (Fig. 6a).

**Fig. 6.**
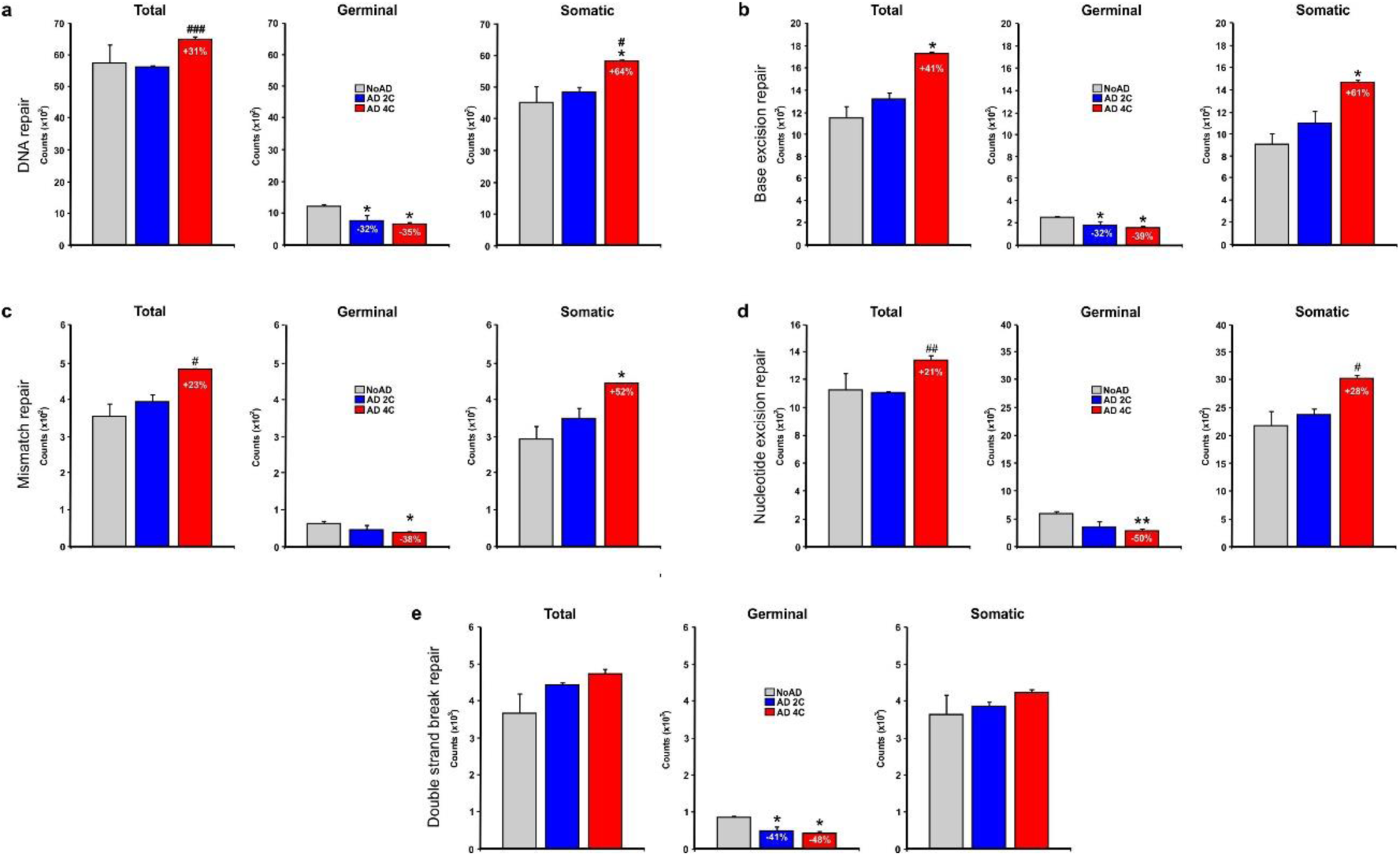
sSNVs counts in DNA repair-associated gene sets of neurons isolated from noAD individuals and AD patients as well as in tetraploid neurons from AD patients. **a** Total (left), germinal (middle) and somatic (right) sSNVs observed in DNA repair genes from noAD diploid neurons (grey), AD diploid neurons (blue) and AD tetraploid neurons (red). **b** As is A for base excision repair genes. **c** As is A for mismatch repair genes. **d** As is A for nucleotide excision repair genes. **e** As is A for double strand break repair genes. *p<0.05, **p<0.01 (comparison NoAD vs AD 4C), ^#^p<0.05, ^##^p<0.01 (comparison NoAD vs AD 4C) (Student’s *t* test).

To explore whether this finding could be extended to all types of genes involved in DNA repair, we focused on genes involved in base excision, mismatch, nucleotide excision, and double strand break repair (see Additional file 2 for a complete list of these genes). This analysis demonstrated that tetraploid neurons displayed a significant increase of total SNVs in all these genes, except those involved in double strand break repair (Fig. 6b-e). For sSNVs, the increase in tetraploid neurons of genes involved in base excision, mismatch, and nucleotide excision was 61 % (Fig. 6b), 52 % (Fig. 6c), and 28 % (Fig. 6d), respectively, while no significant increase of sSNVs was observed in genes regulating double strand breaks (Fig. 6e). As described above for other gene sets, gSNVs were significantly reduced in tetraploid neurons for all DNA repair-associated genes.

Overall, this finding suggests that stochastic mutations in DNA repair genes, due to oxidative stress or other aging-associated mechanisms, facilitates the accumulation of additional somatic mutations due to the lack of capacity to repair the DNA in these cells. This likely facilitates the induction of SNT.

### Absence of enrichment of sSNVs in CR genes

We have previously reported that the expression of oncogenes in the absence of p53 activity facilitates cell cycle reentry in neurons and the survival of the generated hyperploid neurons [3, 4, 56]. Therefore, we hypothesized that tetraploid neurons should contain extra-amount of sSNVs in CR genes, and that the pathogenicity of these mutations should be greater than that observed in other genes. To test these hypotheses, we focused on different proto-oncogenes and tumor suppressor genes (see Additional file 2 for a complete list of these genes). This analysis indicated that the proportion of CR gene-specific sSNVs was significantly higher (23 %) in tetraploid neurons as compared to diploid neurons from both AD patients and NoAD individuals (Fig. 7b). As a control, we focused on the 173 genes that were analyzed by Lodato and collaborators [31], which showed a slightly lower proportion of sSNVs in tetraploid neurons as compared to diploid neurons (20 %) (Fig. 7a). In contrast, the increase of sSNVs observed in CR genes from tetraploid neurons is lower than that observed in genes from the global genome (Fig. 2c). Therefore, the accumulation of cancer-associated gene mutations in tetraploid neurons does not seem to be greater than that observed in control genes.

**Fig. 7.**
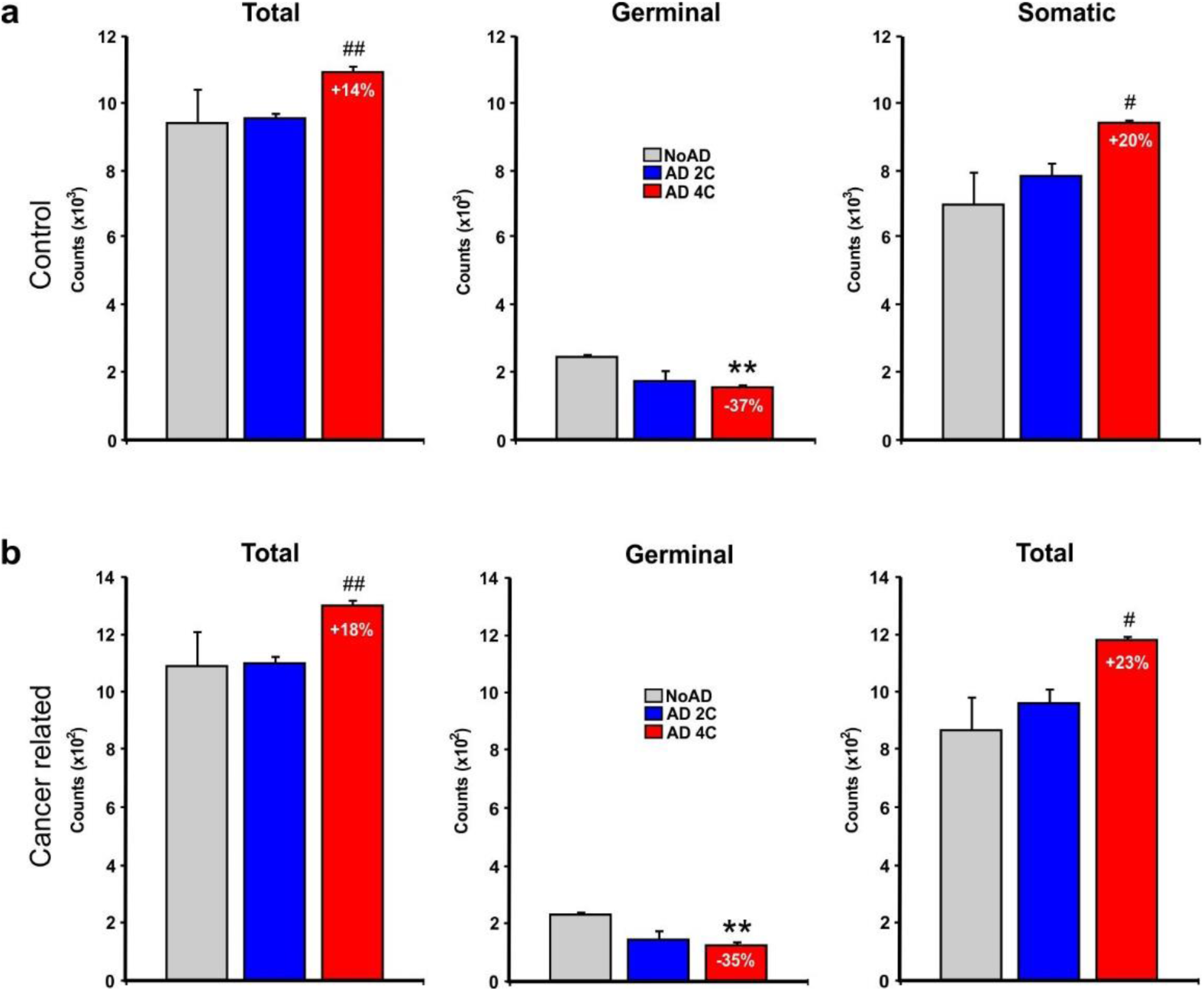
sSNVs counts in CR gene sets of neurons isolated from noAD individuals and AD patients as well as in tetraploid neurons from AD patients. **a** Total (left), germinal (middle) and somatic (right) sSNVs observed in control genes from noAD diploid neurons (grey), AD diploid neurons (blue) and AD tetraploid neurons (red). **b** As is A for CR genes. **p<0.01 (comparison NoAD vs AD 4C), ^#^p<0.05 (comparison NoAD vs AD 4C) (Student’s *t* test).

### CR gene-associated sSNVs show high molecular pathogenicity

To study the expected molecular pathogenicity of the proteins encoded by the genes that accumulate sSNVs in the tetraploid neurons, we used the DG prediction score. This score is calculated from 12 functional prediction algorithms, with 0 being the minimum (less pathogenic variant) and 5 the maximum (most pathogenic variant). Then, the frequency distribution of the prediction score (FDPS) was estimated in both control [31] and CR genes, and the regression slope of the FDPS frequency distribution was used as a measure of the molecular pathogenicity (Additional file 11) (i.e. the greater the slope the stronger the pathogenicity). When the FDPS for the CR gene-specific pool of SNVs was evaluated in tetraploid neurons, a significant enrichment of highly pathogenic SNVs was observed as compared to all sSNVs, evaluated as the statistical difference between the linear regression slope for the tested gene set as compared to the regression slope for the control genes (see [33]) (Table 1). This indicates that CR genes are highly susceptible to mutate to a pathogenic form. This might be involved in SNT, since the absolute proportion of sSNVs is higher in tetraploid neurons (Fig. 2c).

**TABLE 1.** Comparison of the FPDS regression slopes in sSNVs from different gene sets in tetraploid neurons from AD patients. Statistical significance of differences between two linear regression slopes was evaluated using a *t* test based on the slope, standard error, and sample size (López-Sánchez et al., 2021). Reg.: regression. R: Pearson’s r correlation value. DF: Degrees of freedom: 16 for all gene sets.

|  |  | All SNVs (Regression slope: -3.4993; R: 0.901) |  |  |  |  |
| --- | --- | --- | --- | --- | --- | --- |
|  |  | Reg. Slope | R | DF | *t*-value | probability |
| **Generic** | **Walsh** | -4.2915 | 0.8983 | 16 | 0.8324 | 0.4128 |
| **Cancer** | **Cancer** | 0.1867 | 0.2052 | 16 | 5.4579 | 0.0001 |
| **Oxidative stress** | **SOD Activ.** | 1.4815 | 0.3708 | 16 | 3.2900 | 0.0046 |
|  | **Neuron Death** | -1.7427 | 0.4516 | 16 | 1.2264 | 0.2378 |
|  | **Oxidat. Stress** | 0.8768 | 0.5680 | 16 | 4.9882 | 0.0001 |
|  | **Hydrogen Perox.** | -0.1076 | 0.0938 | 16 | 4.7029 | 0.0001 |
|  | **Reactiv. Oxygen** | -0.8786 | 0.1931 | 16 | 1.5003 | 0.1530 |
|  | **Antiox. Activ.** | -2.4929 | 0.6920 | 16 | 0.8439 | 0.4111 |
| **DNA repair** | **DSBR** | -5.0157 | 0.8994 | 16 | 1.4467 | 0.1673 |
|  | **BER** | -0.3989 | 0.2186 | 16 | 3.7771 | 0.0017 |
|  | **MMR** | 2.0602 | 0.6070 | 16 | 5.0228 | 0.0001 |
|  | **NER** | 3.3651 | 0.8201 | 16 | 6.8094 | 4E-06 |
|  | **DNA repair** | -1.9604 | 0.5529 | 16 | 0.7508 | 0.4637 |
| **Senescence** | **Senescence** | -1.0193 | 0.6329 | 16 | 3.3331 | 0.0026 |
| **Neurodegeneration** | **Alzheimer** | -1.3476 | 0.8320 | 16 | 1.1932 | 0.2502 |
|  | **Parkinson** | -3.588 | 0.8628 | 16 | 0.0464 | 0.9635 |

### Variable molecular pathogenicity of other sSNVs

We also evaluated the FDPS of sSNVs in tetraploid neurons from other gene groups by applying the DG prediction score. This analysis indicated that oxidative stress-associated genes showed a variable degree of pathogenicity in tetraploid neurons, with genes involved in SOD activity, oxidative stress, and hydrogen peroxide activity displaying a higher degree of pathogenicity (Additional file 11), which was statistically significant when compared to all sSNVs (Table 1).

DNA repair genes including those involved in base excision, mismatch and nucleotide excision showed a dramatic increase of molecular pathogenicity (Additional file 11), which resulted in significant differences with all sSNVs (Table 1). This contrasted with genes involved in double strand break repair, which in tetraploid neurons showed a low degree of pathogenicity (consistent with the non-significant increase of sSNVs in these genes). The regression slope of these genes (Additional file 11) did not significantly differ from that of control genes (Table 1).

sSNVs from genes involved in senescence also displayed a greater pathogenicity score than those from generic genes in tetraploid neurons (Additional file 11), an effect that was statistically significant (Table 1). This support the notion that the senescent phenotype observed during aging [55] and in AD-affected neurons [12] could be caused by DNA damage followed by cell cycle reactivation and tetraploidy in these cells [21].

Finally, this analysis demonstrated that both AD- and PD-associated genes in tetraploid neurons did not show increased pathogenic capacity when compared with control genes (Additional file 11; Table 1).

Overall, these findings further support the hypothesis that the lack of capacity for DNA repair is what facilitates the accumulation of sSNVs in AD neurons in a context of low capacity to respond to oxidative stress, thus facilitating cancer-like processes in these cells. This mechanism likely participates in the generation of NT in the brain of Alzheimer’s patients.

## Discussion

In this study, we have characterized the somatic mutations present in the genome of tetraploid neurons isolated from the parietal cortex of AD patients, in comparison with those from diploid neurons in this same tissue from both control and paired AD individuals. Different criteria were employed to define whether or not a specific mutation was of somatic origin. We first focused on the frequency at which the mutation was detected. In this regard, previous mutational analyses performed in AD neurons were focused in either bulk whole exome sequencing from several thousands of neurons [46, 47] or single cell whole genome sequencing [27, 36, 37]. The handicap of bulk genome sequencing using genomic DNA from thousands of neurons is the difficulty to detect low-frequency mutations. We have adopted a midway between both approaches as only hundreds of neuronal cells were used for sequencing. This has allowed us to identify low-frequency mutations easily. Since only hundreds of neuronal cell nuclei were whole-genome amplified, those mutations that are present with low frequency were considered to be of somatic origin. The second criterion to define whether or not a defined mutation was somatic was based on whether the mutation is specific for either diploid or tetraploid neurons, instead of being common to both cell types. The finding that most low-frequency mutations are unique for diploid or tetraploid neurons confirmed their somatic nature. Finally, the absence of most low-frequency mutations in existing nucleotide polymorphism databases, known to contain germinal mutations, further confirmed that these low-frequency mutations are of somatic nature.

Two types of somatic mutations were considered in this study, sSNVs and indels, although only the former were enriched in tetraploid neurons. This demonstrates the specificity of the analysis for the identification of somatic mutations in these latter neurons. In general, sSNV mutations were detected in similar chromosomal regions as those of gSNVs, suggesting that mutational hotspots are present in the genome, as previously demonstrated [43]. When the gene-including sequences were studied, sSNVs were found to be enriched in coding exons and promoters. As most sSNVs were missense variants with low stop-gaining and reduced effects on splicing, we can conclude that the mutations enriched in the tetraploid neurons lead to altered protein expression as well as protein malfunction while maintaining intact protein functional domains.

Somatic mutations were previously found in neurons from sporadic AD patients [27, 36, 46]. In our study, no significant increase of sSNVs was detected in diploid neurons from AD patients, in contrast to the study by Li and collaborators [27], which was focused on the whole neuronal population of the prefrontal cortex. Therefore, increased sSNV numbers seems to be specific for SNT. Diploid neurons from control and AD patients were enriched in C>A, C>T and T>C sSNVs, in agreement with the findings of Miller and collaborators [36] and Li and collaborators [27], who found two mutational oxidative stress-associated signatures enriched in these mutations in aged individuals as well as in AD patients.

Other studies dealing with AD-associated mutagenesis have been focused on somatic mutations affecting known AD risk factor genes [7, 44]. Our results confirm the presence of mutations in the AD-risk genes. Nevertheless, mutations associated to PD were also observed to become enriched in the parietal cortex of the AD patients. This indicates that the presence of mutations in risk genes does not necessarily result in the generation of the disease to which these genes are associated.

Somatic mutagenesis takes place randomly and can affect any single gene in the genome, being accumulated with age. In this regard, Lodato and collaborators [31] demonstrated that aging is associated with increased sSNVs in single human neurons from both cerebral cortex and hippocampus. These authors found that cytosine deamination influences patterns of sSNVs, resulting in C>T mutations, which account for most variants in the youngest prefrontal cortex samples. This fraction decreases with age, from above 80 % in the infant cortex to around 50 % in the aged cortex. In contrast, T>C variants increase with age, from less than 5 % in the infant cortex to around 20 % in the aged cortex. We have found that T>C mutations are enriched in tetraploid neurons from parietal cortex, suggesting that these aging-associated, somatic variants participate in pathological NT.

T>C transitions likely represent DNA damage linked to fatty-acid oxidation (lipid peroxidation), triggered by intermediates such as 4-hydroxynonenal (4-HNE) or malondialdehyde (MDA) [11]. This suggests that lipid peroxidation is the major cause of age-associated sSNVs in neurons, which is consistent with the high content of lipids in the nervous system. Therefore, our results suggest that tetraploid neurons have been subjected to high levels of oxidative stress, and this might have triggered SNT, which is consistent with the finding that oxidative stress can induce neuronal polyploidy in Drosophila central nervous system [42]. We cannot rule out that the accumulation of sSNV in genes regulating oxidative stress and DNA repair, and the subsequent SNT could be specific for neurons highly susceptible to oxidative stress [24]. These neurons would thus acquire protection against oxidation due to the genome duplication, as proposed to occur in the brain of Drosophila [42] and of vertebrates [60].

Neurons are postmitotic cells that use the cell cycle machinery to regulate neuronal migration and maturation, and synaptic plasticity [16]. Therefore, it is mandatory for these cells to maintain an exquisite control of this machinery to avoid the orchestration of a functional neuronal cell cycle [15, 56]. Oxidative stress has been postulated to deregulate this control of the cell cycle machinery in neurons, leading to a classical two-hit, oncogenic-like cell cycle [38, 63, 64] that may be at the basis of SNT. This view is consistent with recent studies from our laboratory demonstrating that oncogene expression in the absence of functional p53 can induce cell cycle progression in neurons followed by neuronal hyperploidy [4], and neuronal proliferation when the mitotic check points are pharmacologically abolished [56].

In this study, we provide evidence supporting the two-hit, oncogenic hypothesis initially postulated by Zhu and collaborators [63, 64] for the generation of pathological NT. We have demonstrated that, in tetraploid neurons, sSNVs are highly enriched in genes involved in both oxidative stress response and DNA repair. Therefore, random affection of these genes would potentiate the accumulation of oxidative stress in the affected neurons, which would be exacerbated by the lack of DNA repair in these cells. This would enhance the observed accumulation of sSNVs in other genes, including proto-oncogenes and tumor suppressor genes. This view is consistent with the DNA repair deficiency that is known to occur in neurodegeneration [19, 61]. It is also consistent with the increase of sSNVs in neurons from individuals with Cockayne syndrome or xeroderma pigmentosum, two neurodegenerative phenotypes linked to deficient nucleotide excision repair [31]. In this context, our results indicate that sSNVs with enhanced pathogenicity score are enriched in proto-oncogenes and tumor suppressor genes from tetraploid neurons. This oncogenic-like process would likely participate in the triggering of cell cycle reentry in the AD affected neurons, as previously observed in differentiated neurons [3, 4, 56]. Therefore, we propose a hypothetical model for the generation of tetraploid neurons in the Alzheimer brain based on the mutation of CR genes in diploid neurons due to the inactivation of DNA repair genes, thus facilitating an oncogenic-like cell cycle and SNT (Fig. 8).

**Fig. 8.**
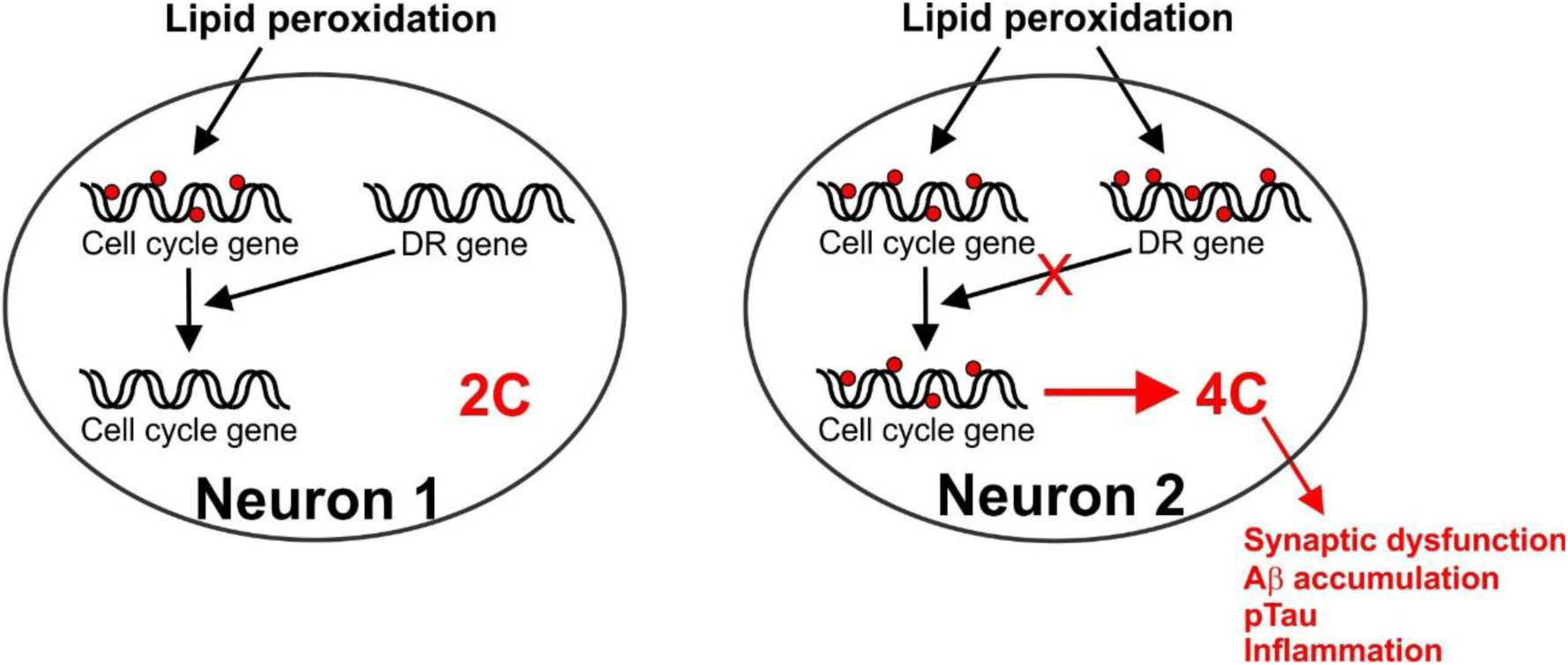
Scheme of the proposed mechanism that triggers the accumulation of sSNVs in diploid neurons, which then become susceptible to become tetraploid. DR: DNA repair.

Our results are in line with the hypothesis that cell cycle reentry in neurons may result from DNA damage response, followed by neuronal senescence in both brain aging and Alzheimer’s disease [57]. In this regard, we have shown that senescence-associated genes show an enhanced pathogenicity score when compared to control genes. This hypothetical equivalence between NT and neuron senescence has profound therapeutic implications, as the multifactorial gene therapy based on E2F4, known to prevent NT among other hallmarks of AD [33, 34, 51], should be considered to possess senolytic capacity.

Anyhow, the understanding of the mechanism leading to neuronal cell cycle reentry and SNT in AD will facilitate the design of effective therapeutic approaches to prevent and, even, cure this devastating neuropathological condition.

## Supporting information

List of patients

Genes that were analyzed per functional category

Relative frequency distribution of the SNVs common to both diploid and tetraploid neurons or specific for either diploid or tetraploid neurons

Low-frequency SNVs in databases containing somatic nucleotide polymorphism

Indels from germinal and somatic origin

Total, germinal, and somatic SNVs located in either coding or non-coding regions

Total, germinal, and somatic SNVs in different chromosomal regions

Total, germinal, and somatic SNVs with different functional features

Specific sSNVs in diploid and tetraploid neurons

sSNVs counts in AD-risk genes

Distribution of pathogenicity scores in sSNVs

## Data Availability

All data produced in the present study are available upon reasonable request to the authors. Whole genome sequencing data will be deposited in the National Center for Biotechnology Information Gene Expression Omnibus (GEO) database before this preprint is published as an article.

## Declarations

## Acknowledgements

The authors thank Eva Sacristán and Jaime Pignatelli for their participation in the bioinformatics analysis.

## Authors’ contributions

NLS performed the experiments and reviewed the manuscript. AR analyzed and interpreted the patient data and reviewed the manuscript. JMF obtained funding and wrote and edited the manuscript. All authors read and approved the final manuscript.

## Availability of data and material

Whole genome sequencing data will be deposited in the National Center for Biotechnology Information Gene Expression Omnibus (GEO) database before this preprint is published as an article.

## Ethics approval and consent to participate

This study has been approved by the Scientific Committee of CIEN Foundation Brain Tissue Bank and the Ethics Committee of *Consejo Superior de Investigaciones Científicas*. Written informed consent for brain removal after death for diagnostic and research purposes was obtained from brain donors and/or next of kin.

## Consent for publication

Written informed consent for research purposes, including publication, was obtained from brain donors and/or next of kin.

## Supplementary Information

**Additional file 1:** List of patients (Excel table).

**Additional file 2:** Genes that were analyzed per functional category (Excel table).

**Additional file 3:** Relative frequency distribution of the SNVs common to both diploid and tetraploid neurons or specific for either diploid or tetraploid neurons (PDF).

**Additional file 4:** Low-frequency SNVs in databases containing somatic nucleotide polymorphism (PDF).

**Additional file 5:** Indels from germinal and somatic origin (PDF).

**Additional file 6:** Total, germinal, and somatic SNVs located in either coding or non-coding regions (PDF).

**Additional file 7:** Total, germinal, and somatic SNVs in different chromosomal regions (PDF).

**Additional file 8:** Total, germinal, and somatic SNVs with different functional features (PDF).

**Additional file 9:** Specific sSNVs in diploid and tetraploid neurons (PDF).

**Additional file 10:** sSNVs counts in AD-risk genes (PDF).

**Additional file 11:** Distribution of pathogenicity scores in sSNVs (PDF).

