## Supplementary material for "Somatic mutations in Alzheimer-associated tetraploid neurons": Relative frequency distribution of the SNVs common to both diploid and tetraploid neurons or specific for either diploid or tetraploid neurons

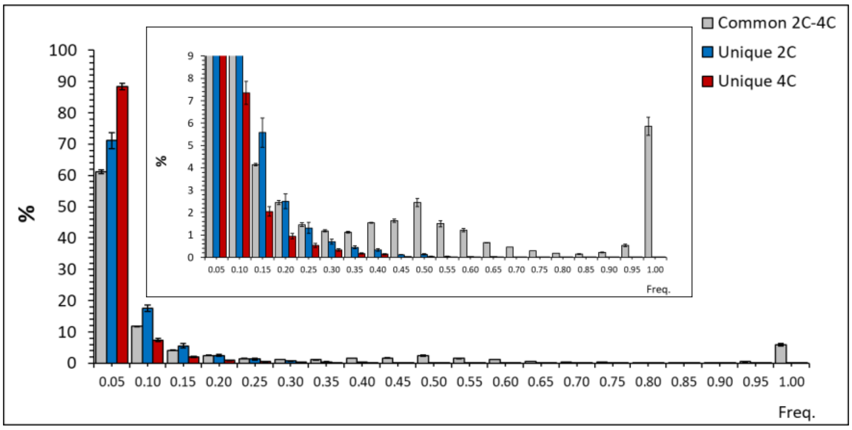


**Additional file 3.** Relative frequency distribution of the SNVs within the same AD individuals (AD409 and AD423), classified as either common to both diploid and tetraploid neurons (i.e. germline variants) or specific for either diploid or tetraploid neurons (i.e. somatic mutations). Box: zoom of the graph indicating percentages ranging from 0 to 9.
