## Supplementary material for "Somatic mutations in Alzheimer-associated tetraploid neurons": Low-frequency SNVs in databases containing somatic nucleotide polymorphism

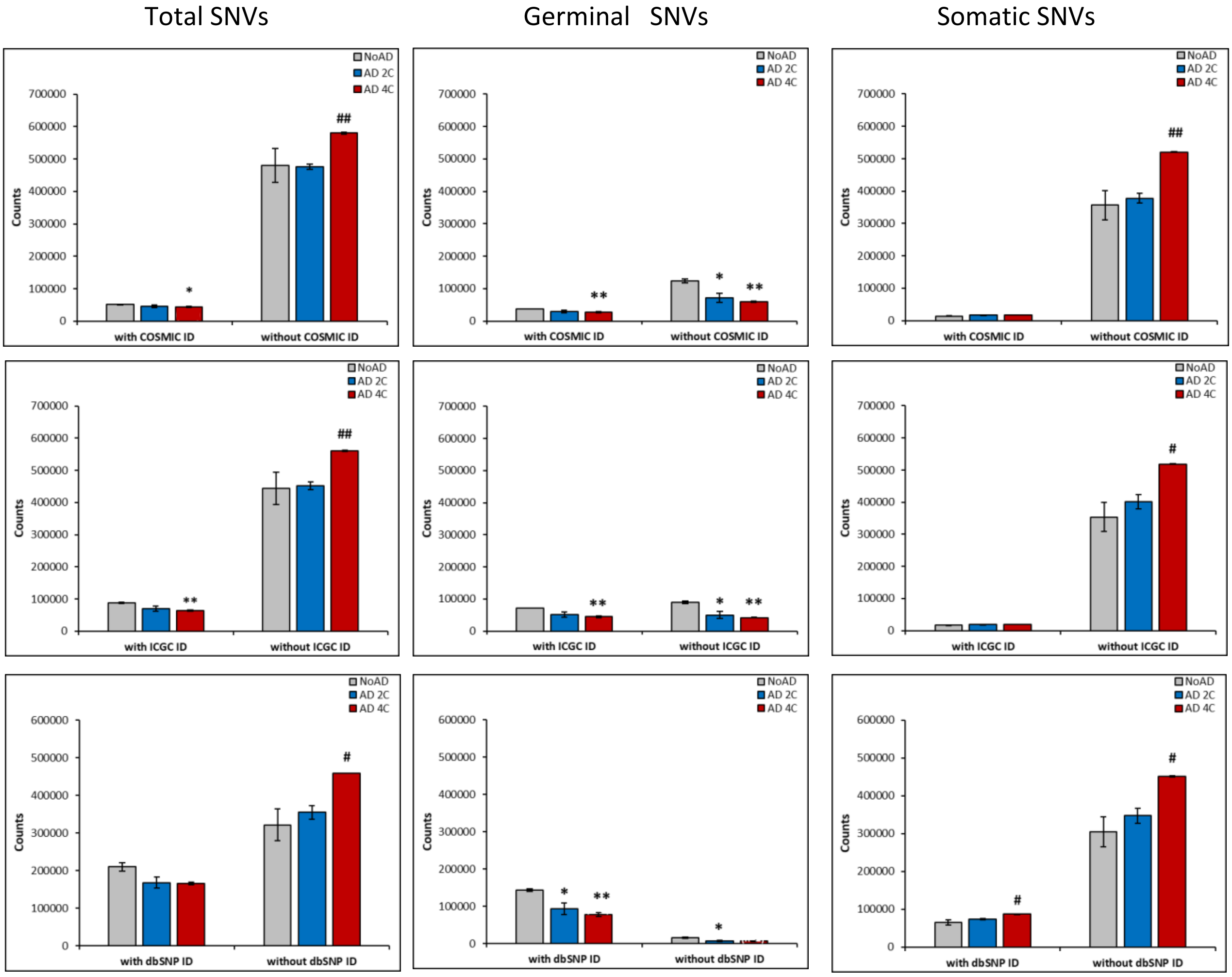


**Additional file 4. Absence of most low-frequency SNVs (i.e. sSNVs) in databases containing somatic nucleotide polymorphism.** Total SNVs, germinal SNVs (i.e. high-frequency SNVs) and somatic SNVs (i.e. low-frequency SNVs) counts with or without identification number (ID) in the catalogue of somatic mutations in cancer (COSMIC), the ICGC data portal, and the reference single nucleotide polymorphism database (dbSNP). *p<0.05, **p<0.01 (comparison NoAD vs AD 4C), #p<0.05, ##p<0.01 (comparison NoAD vs AD 4C) (Student's t test).
