## Supplementary material for "Somatic mutations in Alzheimer-associated tetraploid neurons": Indels from germinal and somatic origin

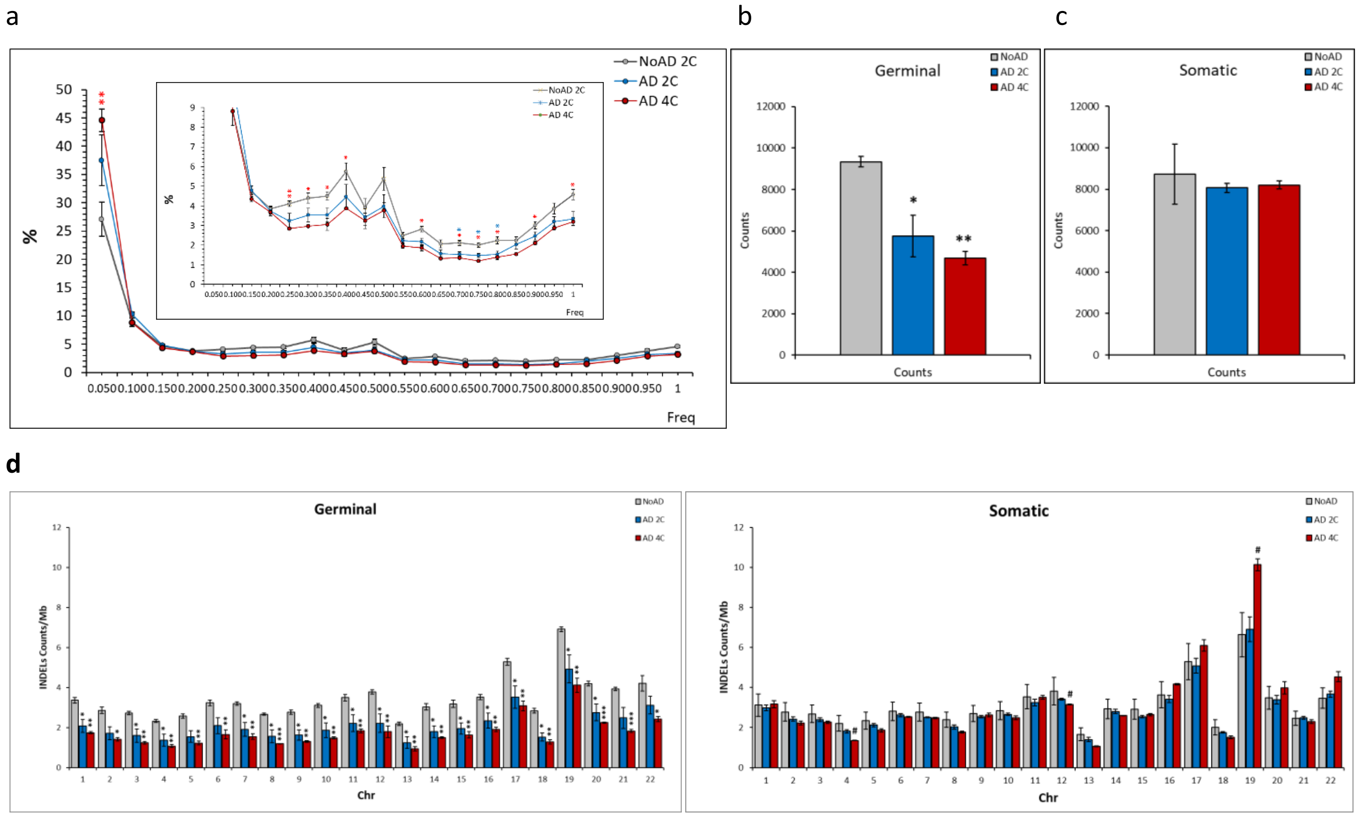


**Additional file 5. Indels from germinal and somatic origin.** a. Relative frequency distribution of the Indels in noAD diploid neurons (grey), AD diploid neurons (blue) and AD tetraploid neurons (red). Dotted line represent the threshold that was used to differentiate indels from germinal (right) and somatic (left) origin. Box: zoom of the graph indicating percentages ranging from 0 to 9. b. Germinal indels observed in noAD diploid neurons (grey), AD diploid neurons (blue) and AD tetraploid neurons (red). c. Somatic indels observed in noAD diploid neurons (grey), AD diploid neurons (blue) and AD tetraploid neurons (red). d. Germinal (left) and somatic (right) indels counts per chromosomal Mb. Numbers identify the chromosomes. *p<0.05, **p<0.01; *p<005 (comparison NoAD vs AD 4C), #p<0.05 (comparison NoAD vs AD 4C) (Student's t test).
