## Supplementary material for "Somatic mutations in Alzheimer-associated tetraploid neurons": Total, germinal, and somatic SNVs located in either coding or non-coding regions

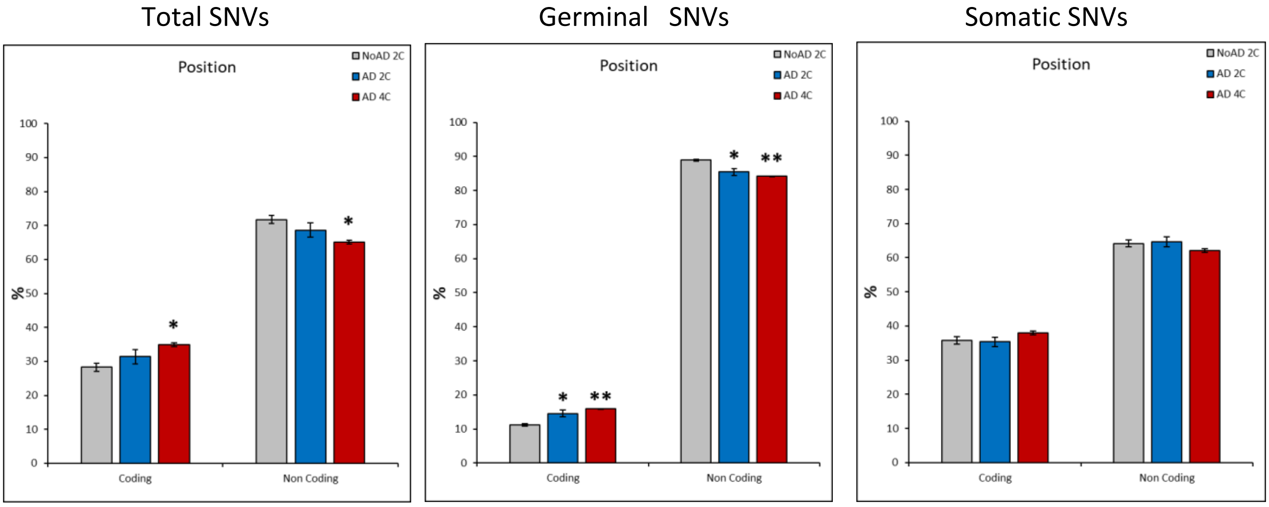


**Additional file 6.** Total SNVs, germinal SNVs (i.e. high-frequency SNVs) and somatic SNVs (i.e. low-frequency SNVs) percentages located in either coding or non-coding regions. *p<0.05, **p<0.01 (comparison NoAD vs AD 4C) (Student's t test).
