## Supplementary material for "Somatic mutations in Alzheimer-associated tetraploid neurons": Total, germinal, and somatic SNVs in different chromosomal regions

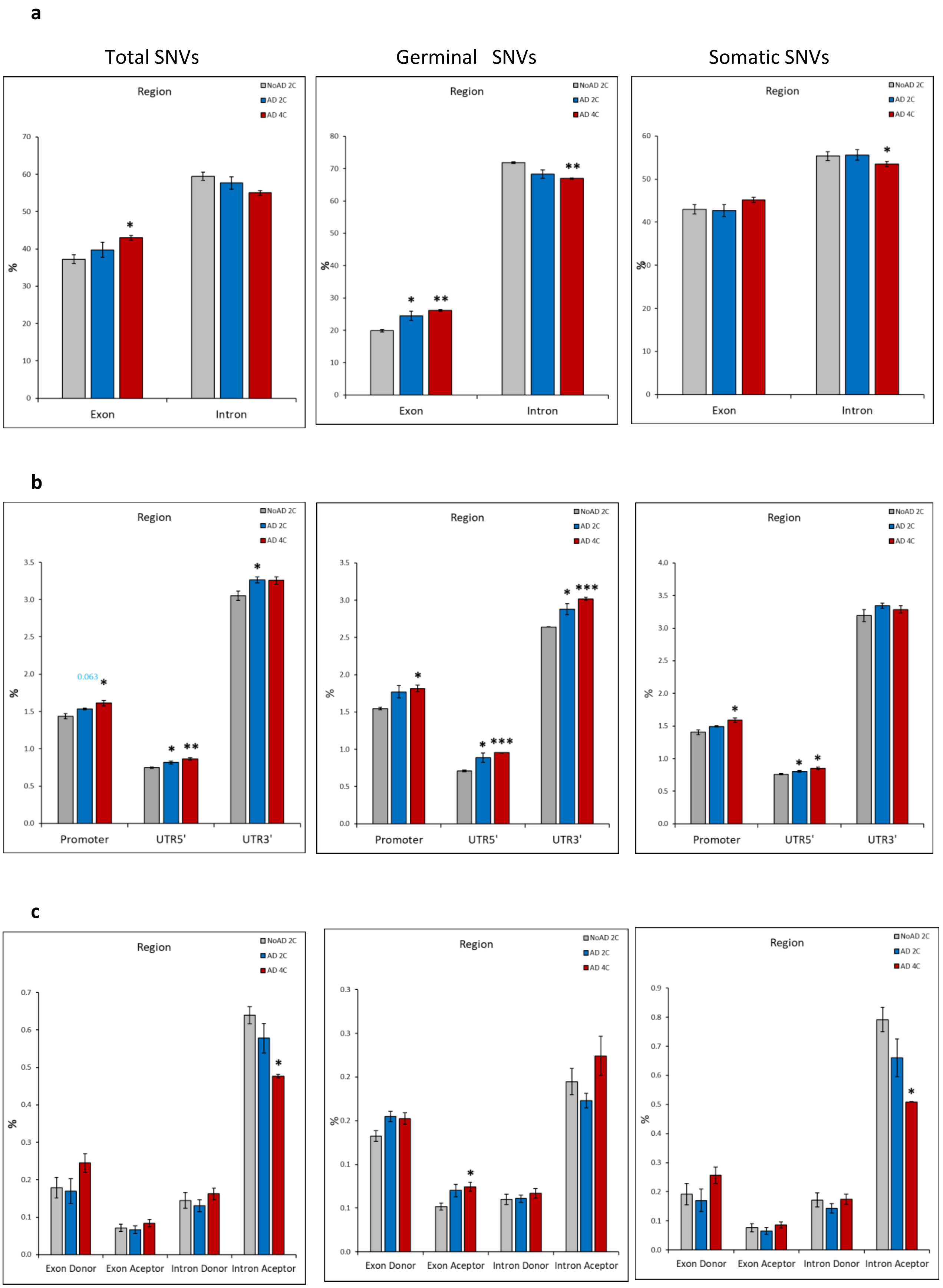


**Additional file 7. Total SNVs, germinal SNVs (gSNVs), and somatic SNVs (sSNVs), in the indicated chromosomal regions from all types of neurons.** a. Percentages of total SNVs, gSNVs (i.e. high-frequency SNVs) and sSNVs (i.e. low-frequency SNVs) located in either exonic or intronic regions from either diploid (2C) or tetraploid (4C) neurons of control and AD patients. b. Percentages of total SNVs, gSNVs and sSNVs located in either promoters, 5’ untranslated regions (UTR5’) or UTR3’ from either diploid (2C) or tetraploid (4C) neurons of control and AD patients. c. Percentages of total SNVs, gSNVs and sSNVs located in either exond donor, exon acceptor, intron donor or intron acceptor regions from either diploid (2C) or tetraploid (4C) neurons of control and AD patients. *$p<0.05$, **$p<0.01$ (comparison NoAD vs AD 4C) (Student’s t test).
