## Supplementary material for "Somatic mutations in Alzheimer-associated tetraploid neurons": Total, germinal, and somatic SNVs with different functional features

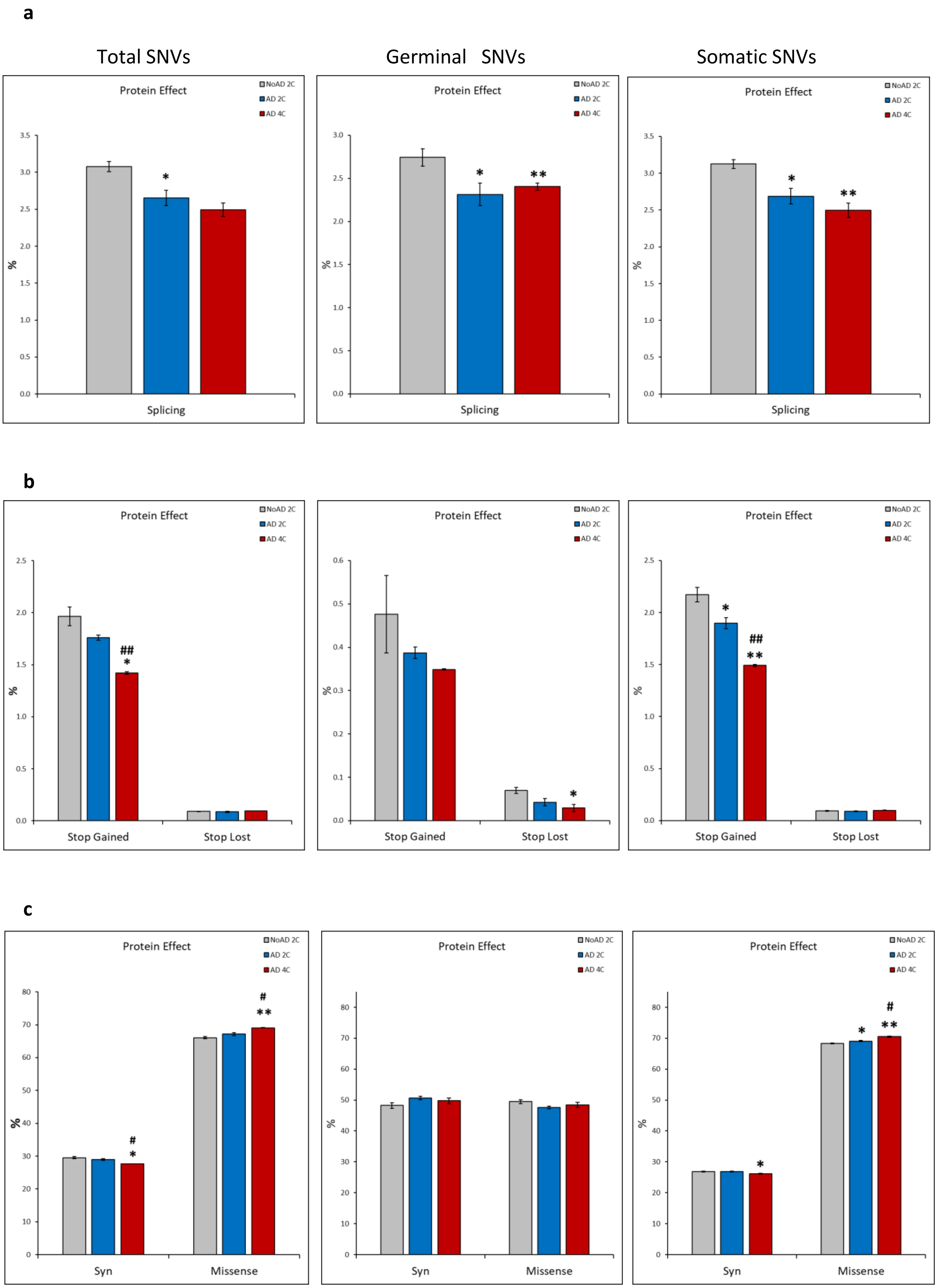


**Additional file 8. Total SNVs, germinal SNVs (gSNVs), and somatic SNVs (sSNVs), with the indicated characteristics in all types of neurons.** a. Percentages of total SNVs, gSNVs (i.e. highfrequency SNVs) and sSNVs (i.e. low-frequency SNVs) located in splicing regions from either diploid (2C) or tetraploid (4C) neurons of control and AD patients. b. Percentages of total SNVs, gSNVs and sSNVs showing either stop codon gaining or loss in diploid (2C) or tetraploid (4C) neurons of control and AD patients. c. Percentages of total SNVs, gSNVs and sSNVs representing synonymous (syn) or missense mutations in either diploid (2C) or tetraploid (4C) neurons of control and AD patients. *p<0.05, **p<0.01 (comparison NoAD vs AD 4C), $^{\#}p<0.05$, $^{\#\#}p<0.01$ (comparison NoAD vs AD 4C) (Student's t test).
