## Supplementary material for "Somatic mutations in Alzheimer-associated tetraploid neurons": Specific sSNVs in diploid and tetraploid neurons

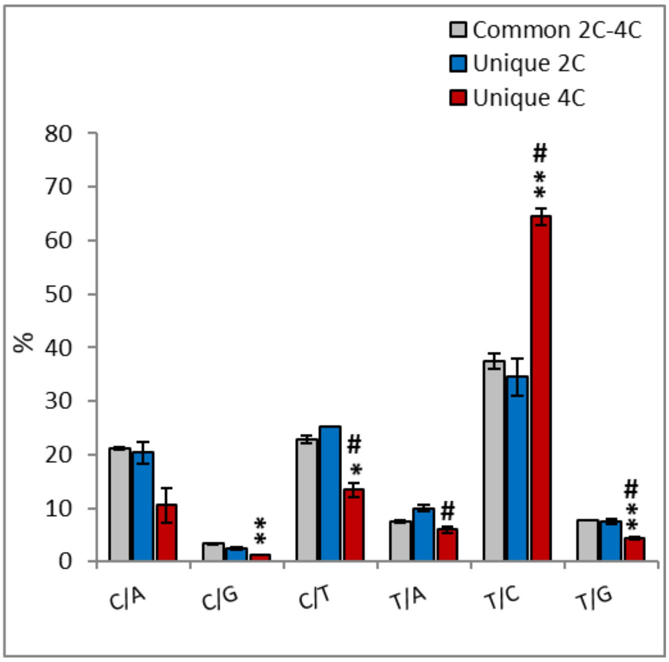


**Additional file 9. Percentage of the identified sSNVs in diploid neurons from noAD individuals and AD patients as well as in tetraploid neurons from AD patients.** A. Percentage of the sSNVs common to diploid and tetraploid neurons (grey) or unique to diploid (blue) or tetraploid neurons (red). C/A (C>A, G>T), C/G (C>G, G>C), C/T (C>T, G>A), T/A (T>A, A>T), T/C (T>C, A>G), T/G (T>G, A>C). *p<0.05, **p<0.01 (comparison NoAD vs AD 4C), #p<0.05 (comparison NoAD vs AD 4C) (Student's t test).
