## Supplementary material for "Somatic mutations in Alzheimer-associated tetraploid neurons": sSNVs counts in AD-risk genes

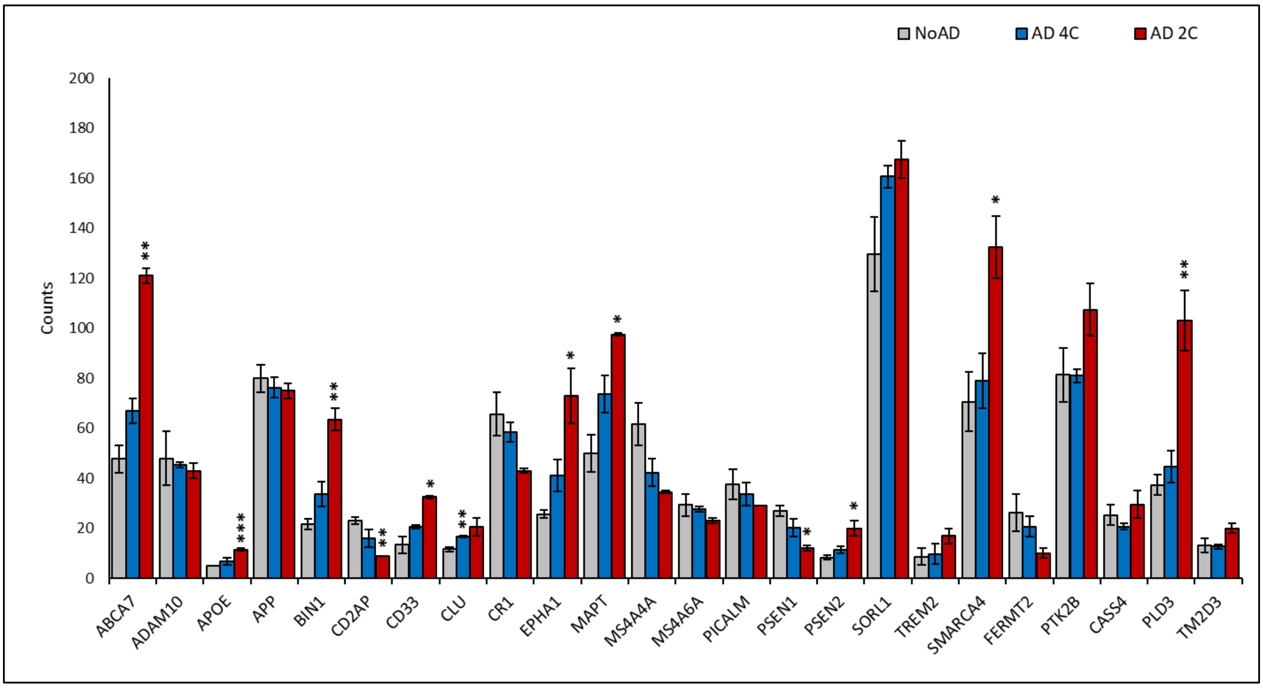


**Additional file 10. sSNVs counts in the indicated AD-risk genes in diploid neurons from noAD individuals or AD patients as well as in tetraploid neurons from AD patients.** *p<0.05, **p<0.01, **p<0.001 (comparison NoAD vs AD 4C) (Student's t test).
