## Supplementary material for "Somatic mutations in Alzheimer-associated tetraploid neurons": Distribution of pathogenicity scores in sSNVs

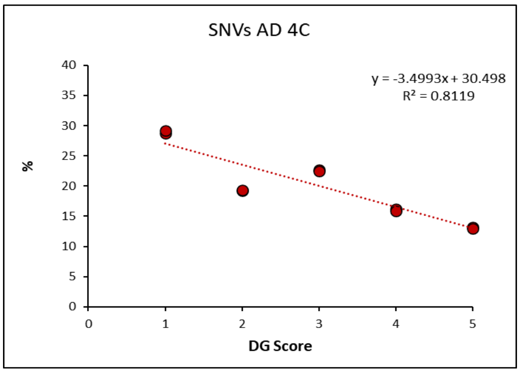
SNVs AD 4C
y = -3.4993x + 30.498
R² = 0.8119
%
DG Score

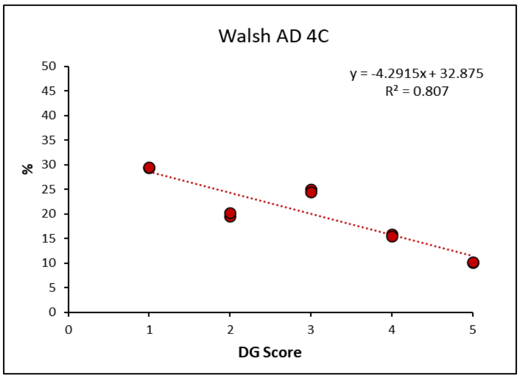
Walsh AD 4C
y = -4.2915x + 32.875
R² = 0.807
%
DG Score

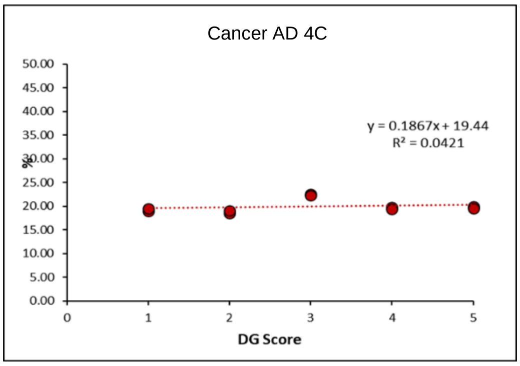
Cancer AD 4C
y = 0.1867x + 19.44
R² = 0.0421
%
DG Score

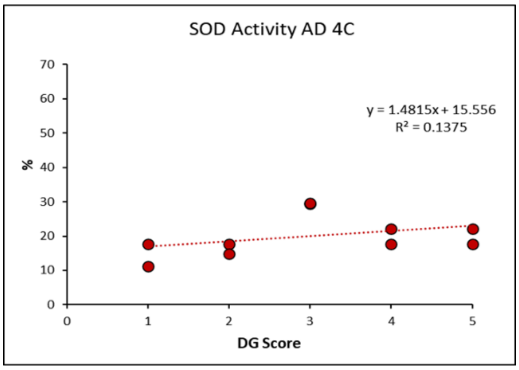
SOD Activity AD 4C
y = 1.4815x + 15.556
R² = 0.1375
%
DG Score

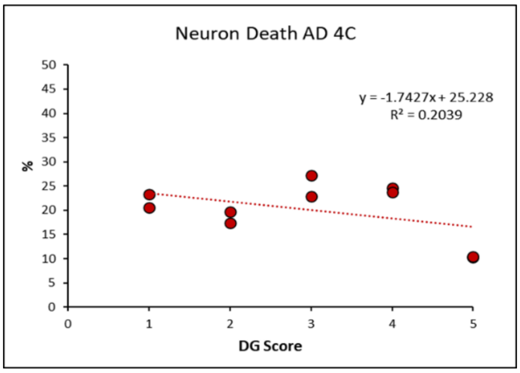
Neuron Death AD 4C
y = -1.7427x + 25.228
R² = 0.2039
%
DG Score

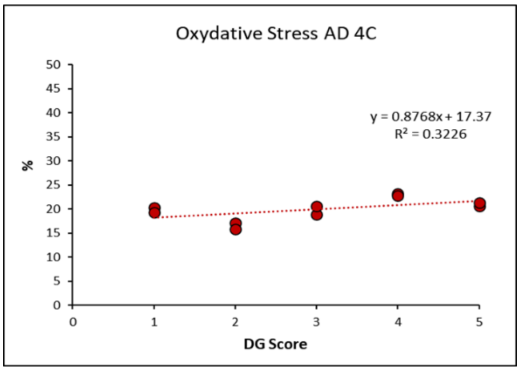
Oxydative Stress AD 4C
y = 0.8768x + 17.37
R² = 0.3226
%
DG Score

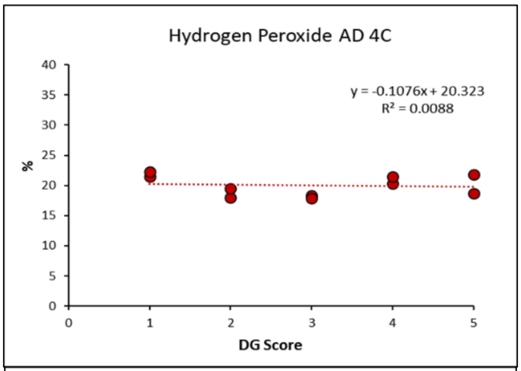
Hydrogen Peroxide AD 4C
y = -0.1076x + 20.323
R² = 0.0088
%
DG Score

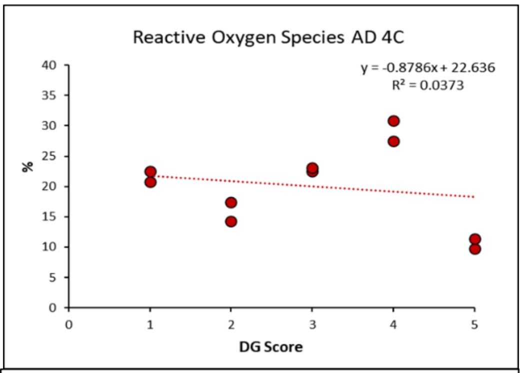
Reactive Oxygen Species AD 4C
y = -0.8786x + 22.636
R² = 0.0373
%
DG Score

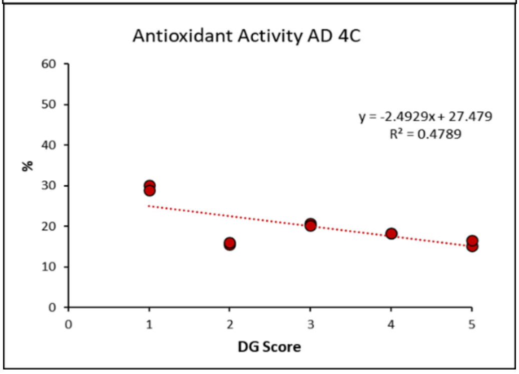
Antioxidant Activity AD 4C
y = -2.4929x + 27.479
R² = 0.4789
%
DG Score

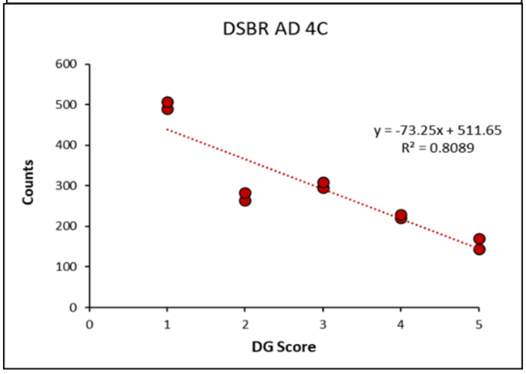
DSBR AD 4C
y = -73.25x + 511.65
R² = 0.8089
Counts
DG Score

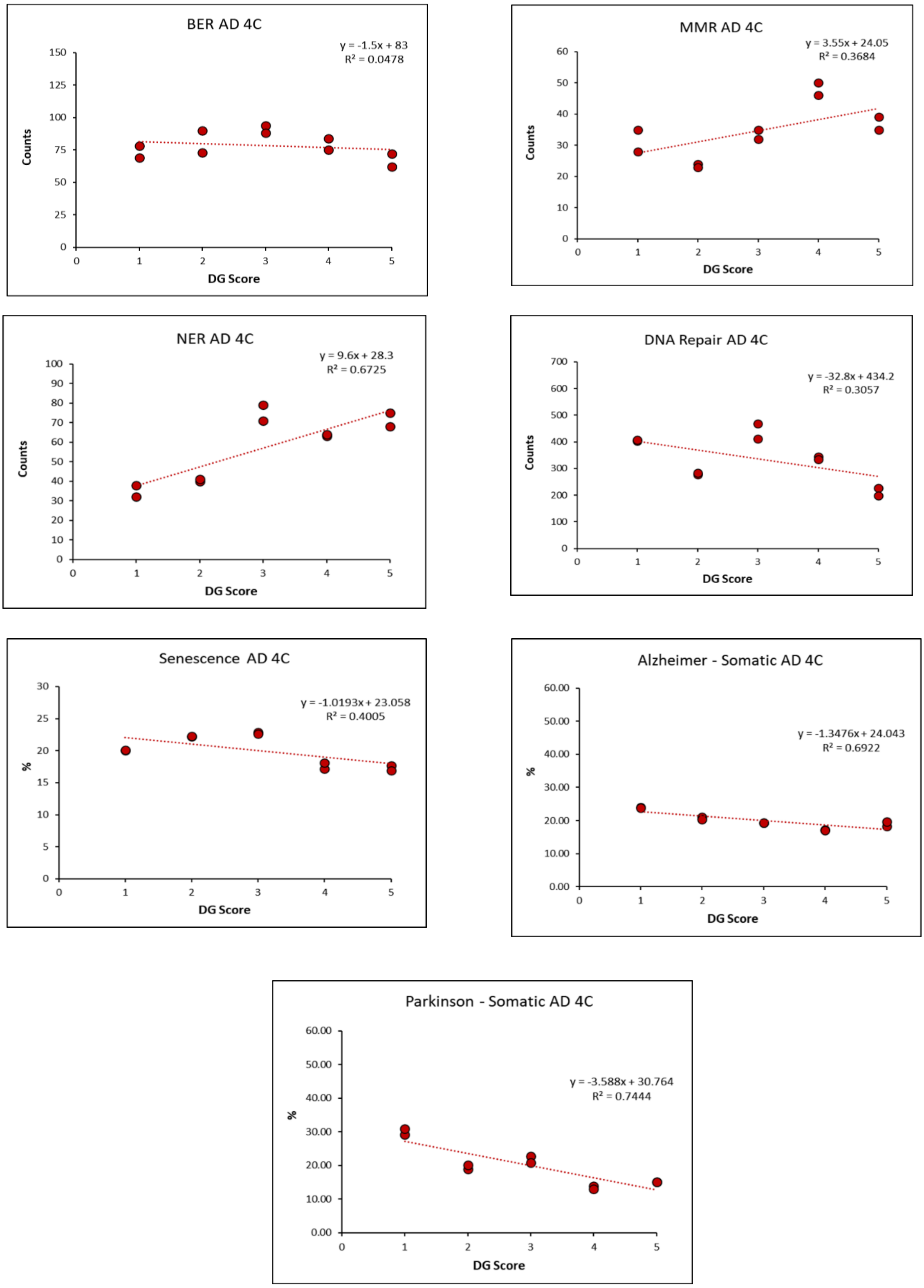


**Additional file 11.** Distribution of pathogenicity scores in sSNVs from the indicated gene sets of tetraploid neurons isolated from AD patients. Regression curves and coefficient of regression ($R^2$) for the relative frequency distribution of the pathogenicity of the sSNVs is shown.
